# A patient-centered comparative effectiveness research study of culturally appropriate options for diabetes self-management

**DOI:** 10.1101/2023.01.31.23285236

**Authors:** Janet Page-Reeves, Cristina Murray-Krezan, Mark R. Burge, Shiraz I. Mishra, Lidia Regino, Molly Bleecker, Daniel Perez, Hannah Cole McGrew, Elaine L. Bearer, Erik Erhardt

## Abstract

This project compared the effectiveness of two evidence-based models of culturally competent diabetes health promotion: *The Diabetes Self-Management Support Empowerment Model* (DSMS), and *The Chronic Care Model* (CCM). Our primary outcome was improvement in patient capacity for diabetes self-management as measured by the Diabetes Knowledge Questionnaire (DKQ) and the Patient Activation Measure (PAM). Our secondary outcome was patient success at diabetes self-management as measured by improvement in A1c, depression sores using the PHQ-9, and Body Mass Index (BMI). We also gathered data on the cultural competence of the program using the Consumer Assessment of Healthcare *Providers and Systems Cultural Competence Set* (CAHPS-CC). We compared patient outcomes in two existing sites in Albuquerque, New Mexico that serve a large population of Latino diabetes patients from low-income households. Participants were enrolled as dyads—a patient participant (n=226) and a social support participant (n=226). Outcomes over time and by program were analyzed using longitudinal linear mixed modeling, adjusted for patient participant demographic characteristics and other potential confounding covariates. Secondary outcomes were also adjusted for potential confounders. Interactions with both time and program helped to assess outcomes. This study did not find a difference between the two sites with respect to the primary outcome measures and only one of the three secondary outcomes showed differential results. The main difference between programs was that depression decreased more for CCM than for DSMS. An exploratory, subgroup analysis revealed that at CCM, patient participants with a very high A1c (>10) demonstrated a clinically meaningful decrease. However, given the higher cultural competence rating for the CCM, statistically significant improvement in depression, and the importance of social support to the patients, results suggest that a culturally and contextually situated diabetes self-management and education program design may deliver benefit for patients, especially for patients with higher A1c levels.

## Introduction

### Overview

Here we report results from a non-randomized, pragmatic, quasi-experimental, patient-engaged, comparative effectiveness research (CER) study [1] conducted by researchers at [BLINDED] comparing two distinct evidence-based models for culturally competent diabetes self-management and education programming. We compared outcomes at two sites in Albuquerque, New Mexico, serving a large Latinx patient population from low-income households. Data reported here were gathered at baseline between February 2016 and March 2020. The overall study protocol is described in detail elsewhere [1]. Data collection involved interviews, focus groups, surveys, and assessments of each program, and testing of patient participants for A1c, depression, and Body Mass Index (BMI). Survey responses, blood samples, and height/weight measurements were gathered at four time points (baseline, 3-monts, 6-months, and 12-months). The [BLINDED] Human Research Review Committee/Institutional Review Board (HRRC/IRB # 16-303) approved all aspects of the research protocol.

### Background

#### The specter of uncontrolled diabetes

Diabetes is among the Institute of Medicine’s top 25 national priorities for Comparative Effectiveness Research (CER) [2]. More than thirty-seven million people or 11.3% of the U.S. population have Type 2 diabetes [3]. While statistics this large can seem remote and impersonal, each patient member of our project team has life-altering personal experience with diabetes. Not only do they have diabetes or pre-diabetes themselves, they also have family members or multiple family members with diabetes. They report that diabetes is so common in the Latinx community that people just assume that they will get it, and if they are diagnosed with diabetes, that diabetes is a death sentence about which there is nothing they can do. So, most of the time, they do nothing. Our patient team members fear diabetes not only for themselves and their adult family members, but also for the future that awaits their children growing up with the specter of diabetes but without the knowledge, capacity, or skills to take control of their own health destiny. Our project sought to disrupt this fatalistic dynamic of despair. As such, our partners, who are Latinx patients from low-income households, their family members, and healthcare providers who serve this population of patients recognize effective diabetes self-management as a matter of life and death.

#### Diabetes health disparities

Although diabetes is a national health crisis, risk is not the same for everybody Individuals from minority and ethnic populations and those with low-income status are at significantly higher risk [4]. Latinx adults are 70% more likely than non-Latinx white adults to be diagnosed with diabetes by a physician [5] and the risk of developing diabetes over the lifespan for Latinxs is 50% versus 40% for the overall U.S. population. According to an analysis of data from the U.S. National Longitudinal Mortality Study, Latinxs are also 28% more likely to die from diabetes, with Mexican Americans, representing 33.5 million people or 61.4% of U.S. Latinxs,)] 50% more at risk [6]. Rates for diabetes diagnoses (11.9%) and the diabetes death rate (45.9 per 100,000) for Latinxs are both more than twice those for non-Hispanic whites (5.3% and 22.5) [7,8]. Not surprisingly, a national poll by Harvard University, National Public Radio (NPR), and the Robert Wood Johnson Foundation found that diabetes is the top health concern for Latinx families [9].

Similarly, poverty has an impact on diabetes risk. Research shows that individuals from low-income communities experience higher rates of diabetes [10–13]. Analysis of National Health Interview Survey data found that the *“*greatest disparities [for diabetes risk] were experienced by the groups who had the lowest level of education, were living below the Federal Poverty Level (FPL), or both [4].” This is a troubling concern for Latinxs given the high level of Latinx poverty (15.7%) [11]. In New Mexico, where Latinxs, who make up 47% of the population, have a poverty rate of 20.8% [14], ethnicity and poverty both play a significant role in diabetes health and health disparities. New Mexico is the second-poorest state in the nation, with poverty among Latinxs (24% for 18-64 year-olds and 37% for 17-and-under) significantly higher than among non-Hispanic whites (12% and 13%) [15]. Individuals in New Mexico from low-income households are nearly three times more likely to be diagnosed with diabetes (14%) than individuals from households making more than $50,000 (5.2%) [3], meaning that given the Latinx poverty rate, diabetes risk for individuals from low-income Latinx households is disproportionately high.

#### Gaps in evidence

Biomedical approaches to diabetes care are well-established, but pharmacologic therapies are often extremely costly, may have problematic health side effects, do not always result in the intended improvement in patients’ diabetes health, and involve hard-to-follow regimens given social and environmental barriers faced by low-income patients. Instead, health guidelines emphasize the important role of patient self-care over narrow reliance on medical treatments for reducing the health impact of diabetes and improving diabetes health outcomes. The *Guide to Community Preventive Services* instructs individuals to engage in lifestyle changes based on combined diet and physical activity improvements as the best way to prevent and manage Type 2 diabetes [16]. The *Michigan Quality Improvement Consortium Guidelines for Management of Diabetes Mellitus* recommend that individuals be given “comprehensive diabetes self-management education.” [17]. *Recommended Lifestyle and Self-management Guidelines from the American Diabetes Association* discuss the importance of individualized education, monitoring, and counseling [18]. Individuals can self-manage their diabetes or prevent pre-diabetes from becoming full diabetes through daily physical activity, a healthy diet, minimizing stress, and for those with full diabetes, regular glucose self-monitoring [19]. But these are not things that can happen in the clinic or via prescription; they are things that patients must do to care for themselves every day.

Despite clear and consistent guidelines, diabetes health outcomes are not improving [19]. The guidelines do not provide a roadmap for getting individuals to embrace necessary self-care practices. However, systematic reviews have repeatedly demonstrated that culturally competent health promotion approaches that account for a patient’s culture and the social context of poverty can be vital to improving health outcomes [20–28]. In particular, culturally competent self-management interventions have been shown to significantly improve glycemic control and behaviors related to diet and physical activity, and increase diabetes-related knowledge. As a result, “cultural competence” has become prominent in diabetes health promotion. Various models have been developed to create “culturally competent” diabetes self-management programming [20–28]. Yet, there is no agreement on what “cultural competence” actually means or entails. Because of a continued emphasis on individual behavior in approaches to diabetes health promotion, the design of self-management programming does not always create cultural competence in a way that makes sense in the context of patients’ lives or improves their health.

#### Promising practices

Different models are being used to make diabetes self-management programs culturally competent. However, this variation creates uncertainty for a patient with diabetes who does not understand that programs can differ significantly, how they differ, or which programs offer them the best option. For Latinx patients from low-income households, it is not clear which type of culturally competent self-management programming most effectively integrates their culture and accommodates their socio-economic circumstances in a way to best improve their diabetes health. This study addressed this gap by using issues identified as important by patients to design measures for directly comparing different evidence-based models for culturally competent diabetes self-management health promotion being implemented by programs that are currently available to Latinx patients from low-income households in Albuquerque, New Mexico. Our patient-engaged preliminary research suggested that the results of this study could offer significant benefit to patients trying to find support for developing the knowledge and capacity to self-manage their diabetes [29,30]. Patient stakeholders recognized the imperative of everyday diabetes self-care strategies, but lacked the knowledge and capacity to successfully adopt the changes outlined in the guidelines, thus improving their Hemoglobin A1c (glycosylated hemoglobin) and successfully controlling their diabetes. Improving models and approaches for diabetes self-care health promotion is critical to the health of our Advisors, their adult family members, and their children.

### Approach

#### Community Engaged Design

We used a community-engaged research approach with the engagement of, and participation by, diverse patient stakeholders, including Latinx diabetes patients and their social supports, Latinx community health workers (CHWs), Latinx diabetes educators [31–34], and partner community agencies serving Latinx clients. A patient advisory board identified the research question and contributed to the design and implementation of the study. Patient stakeholders participated in all aspects of the research. The community co-principal investigator (Co-PI), the project coordinator, the primary research site director, the research manager, and three data collectors were from the population of study [31,34]. We convened a 10-member

Patient Advisory Board (PAB) of patients, individuals who provide care or social support to a person(s) with diabetes (hereafter, “social supports”), and researchers. Patients and patient stakeholders participated in the study across the continuum of engagement, including shared leadership, collaboration, consultation, and input at every stage.

#### Rationale

The idea for this study came from our patient partners. In 2009, a CHW at One Hope Centro de Vida Health Center (referenced hereafter as One Hope) who knew PI Page-Reeves asked her for help because the CHW saw a rising problem of diabetes in the Latinx community that was not being addressed, and she felt that patients did not really understand their diabetes or know what to do when they were diagnosed. We obtained pilot funding through [BLINDED] to assess the problem’s dimensions and obtain community input regarding diabetes health and ideas for prevention. We mapped local Geographic Information System (GIS) data and conducted a survey and blood analysis for A1c with 100 people, conducted interviews with key community stakeholders, and held a series of focus groups with patients and social supports. Our PAB participated in designing focus group/interview questions and interpreting findings. Results from the pilot [29,30] provided a roadmap for a CCM diabetes self-management initiative at One Hope. This laid the foundation for us to receive funding from the Patient-Centered Outcomes Research Institute (PCORI) to engage patients, social supports, stakeholders from different diabetes programs, and university researchers in designing this study.

We provided a respectful and culturally appropriate environment for PAB meetings [33]. Our PAB included individuals who were uncomfortable speaking in English or did not understand English. PAB meetings were conducted in Spanish as the default language. If we brought in content experts or researchers who did not speak Spanish, we provided them with an English translation, but we still held the meetings in Spanish to acknowledge patients as at the core of the process. During the planning process, we provided trainings to help our PAB members develop their capacity to participate in and contribute to the research. We held PAB meetings at One Hope clinic, an accessible location for PAB members that does not have the challenges of parking that exist at the university.

### Models for comparison

#### Local context for each model

The two models being compared were the Diabetes Self-Management Support Empowerment Model (DSMS) and [35] the Chronic Care Model (CCM) [28,36]. Each program serves a large population of Latinx patients from low-income households in Albuquerque, New Mexico, and employs a distinct evidence-based approach to create program cultural competence. Patient participants were recruited from both programs. One program, the Center for Diabetes Education at the University of New Mexico Hospital (CDE-UNMH), is based at a university hospital and uses the DSMS [35]. The other, One Hope, is based at a community clinic operated by a faith-based nonprofit and uses the CCM approach [28,36].

#### The Diabetes Self-Management Support Empowerment Model (DSMS)

The DSMS is a patient-centered, theoretically based educational framework that follows National Standards for Diabetes Self-Management Education [35], is certified by the American Diabetes Association [37], and is accredited by the American Association of Diabetes Educators (AADE) [37]. The DSMS combines a series of clinically informed group didactic sessions that use a patient self-determination approach to empower patients to take control of their own diabetes health with follow-up support to sustain self-management gains achieved during the sessions. The AADE requires that educators acquire proficiency in culturally competent supportive care across the lifespan as one of five domains for certification so that educators can be informed about and aware of specific challenges that might accrue in the patient’s diabetes self-management experience. This program represents the gold standard for diabetes self-management education, focusing on changing eating and physical activity behaviors, self-monitoring, risk reduction, and stress management. The CDE-UNMH program uses the DSMS group education approach. Patients attend a six-week group instructional session with nine hours of class plus a one-on-one follow-up with a certified diabetes educator to provide individualized support by creating a customized education plan. The group sessions have discussions supported by didactic conversation “maps” where the facilitator guides but does not control the conversation based on session thematic goals. Patients then complete self-assessment forms. This format is the foundation of the DSMS Model for creating patient empowerment and program cultural competence.

#### The Chronic Care Model (CCM)

The CCM is “a systematic approach to restructuring medical care to create partnerships between health systems and communities” [28,36] by addressing not only the medical but also the cultural and linguistic needs of patients through the inclusion of cultural competence in the delivery system design [28].The CCM involves six synergistic domains: *1.)* Improved access to care, *2.)* Patient self-management support, *3.)* Patient decision support, *4.)* Care coordination, *5.)* Integrated health information systems, and *6.)* Access to community resources. The use of the CCM framework has yielded significant results in treating diabetes and is being used widely in chronic disease management [36]. To create a holistic care regime, the CCM focuses on addressing social determinants of health by meeting patients’ medical, cultural, and linguistic needs through integrating cultural norms and social relationships from the patient population into program design [24].

The *One Hope Program* is based on the CCM and is designed to address the specific needs of Latinx patients from low-income households [25,30] by creating comprehensive, integrated, wrap-around services focused on culturally competent care [24]. One Hope emphasizes Spanish as the language for service provision [29,30] and access to care regardless of ability to pay. The One Hope facility provides a physical environment that reflects patients’ lifestyle and economic capacity to make them feel comfortable and that they “belong” (in contrast to more clinical, corporate, or academic medical settings). One Hope is a community-run clinic with a director and staff who are members of the community and who are culturally and economically similar to the patients they serve, reducing the hierarchical power relationship that generally exists between patients and providers. This approach is evident in the way that doctors at One Hope share decision-making by engaging the patient and their family members in creating a plan for diabetes self-management.

In addition, patients, caregivers, and family members participate in a variety of program activities including cooking and nutrition workshops, *zumba* classes, and *citas compartidas* sessions (“shared appointments”) [38,39]. These shared appointment sessions allow patients, social supports, and family members to share their stories and experiences in a peer support setting with facilitation by medically trained providers. But providers also “co-learn” from the patients [25,40,41]. Through shared decision-making and shared appointments, providers learn about the realities of patients’ lives and their daily struggles at a level beyond the interaction normally occurring in a clinic. This helps the provider to be culturally competent by understanding diabetes from the perspective of the patient. Sharing experiences with peers and providers and including family members in activities offers a different level of social support for the patient by creating an enhanced feeling of intimacy and inclusion within the program.

Innovative *salidas* (exit interviews), conducted routinely with all patients by a bilingual health navigator, ensures that the patient understands and feels capable of implementing a doctor’s instructions, and integrates care delivery by allowing the health navigator to communicate details of patient status back to the provider [29].

### Research question

#### Development of the research question

> “I have diabetes, but I am not just a patient. I am a person. I have cultural values and concrete realities that shape my everyday life. Both need to be considered for me to be able to feel that my care is making me well and to make it more likely that I can control my A1c. With this in mind, which of two self-management programs is the most culturally and contextually appropriate option for me <u>to take the best care of</u> <u>myself</u> in relation to my diabetes?”^[Translation from Spanish]^

We co-developed this research question with our PAB. Specifically, they were concerned about the failure of diabetes self-management programming to account for important dimensions of Latinx culture or the social context created by poverty. At our PAB meetings, patients and social supports discussed these issues with us extensively and with emotion. What they had to say supported what we heard in previous conversations with patients and community members, and in our preliminary research [29,30]. Patients, social supports and community members reported: a.) A lack of cultural competence on the part of providers, b.) A lack of programming in Spanish, c.) Failure of program design to understand or accommodate the dynamics of Latinx culture related to core values prioritizing the importance of social relationships and the need to avoid personal conflict [41,42]. d.) Poor program accommodation of the fact that patients lack resources, and [40] e.) A lack of attention to the extent to which poverty results in low diabetes health literacy, low capacity to deal with chronic disease, and high stress [40]. All of these factors influence patients’ ability to comply with recommendations regarding drugs, diet, and physical activity to self-manage their diabetes. This question guided the research.

#### Theoretical framework

Trickett proposes that cultural competence entails integrating components of an intervention “into the local expression of culture as reflected in the multiple levels of the ecological context” [43,44]. Rather than merely “tailoring” an existing intervention to target a specific context or population (for example, by offering recipes for healthy meals using Latinx cuisine or providing educational materials in Spanish), he emphasizes the need for interventions to be “situated” to fit synergistically within broader community dynamics (culture and socio-economic context). Following Trickett, and reflecting input from our patient partners, we hypothesized that getting people to adopt lifestyle and behavior changes outlined in guidelines for diabetes self-management requires positively leveraging the cultural values and accommodating the socio-economic circumstances of a patient population in a way that creates synergy with specific social dynamics that define patients’ everyday lives [29,30] (Fig 1).

Therefore, our overarching study hypothesis was that diabetes self-management programs are most successful if their design is culturally and contextually “situated.”

Fig 1. **Diagram of relationships between study elements**. A1c levels is used as a biomarker of serum glucose, and indiator of diabetes. A1c, glycated hemoglobin A1c; BMI, body mass index.

#### Specific aims

This study had three specific aims:

> **Aim #1.** Measure and compare the improvement in patient <u>capacity</u> for diabetes self-management as indicated by improvement in diabetes knowledge and patient activation.
>
> **Aim #2.** Measure and compare patient <u>success</u> at diabetes self-management as indicated by improvement in A1c, depression index score, and body mass index (BMI).
>
> **Aim #3.** Characterize the ways that two distinct culturally competent diabetes self-management programs <u>interface</u> with patient culture and socioeconomic context.

### Outcome measures

While changes in diet or levels of physical activity are commonly understood as measures of diabetes health [19], our PAB told us that acquiring the capacity for diabetes self-management must occur first. They said there is too much focus on diet and physical activity when other things are more important for getting people to the point where they can take care of themselves. They said that if they do not understand their diabetes, they cannot even begin to self-manage their condition. But beyond knowledge, they said that it is important for researchers to determine what helps patients move from knowing to taking action. They saw “capacity”—our primary patient-reported outcome—as comprised of this combination of knowledge and ability to take action.

#### Improved patient capacity for diabetes self-management

For our primary outcome, we identified two validated and reliable instruments available in both English and Spanish. Capacity for diabetes self-management was measured using the Diabetes Knowledge Questionnaire (DKQ) and the Patient Activation Measure (PAM).

1. Diabetes knowledge was measured using the *DKQ summed score*. <u>Hypothesis</u>: The *Chronic Care Model* (CCM) model would result in a larger increase in DKQ summed scores from baseline to 6 months as compared to the *Diabetes Self-Management Support Empowerment Model* DSMS. Previously published studies evaluating culturally competent diabetes management programs report meaningful changes in DKQ summed scores with Cohen’s *f* effect sizes of 0.03 to 0.16 in studies ranging in sample sizes per arm from 10 to 189 [45–48].
2. Patient activation was measured using the *PAM-10 raw score*. <u>Hypothesis</u>: The CCM model would result in a larger increase in PAM-10 raw scores from baseline to 6 months as compared to the DSMS. Previously published studies evaluating culturally competent diabetes management programs report changes in PAM-10 raw scores with meaningful Cohen’s *f* effect sizes of 0.01 to 0.16 in studies ranging in sample size per arm from 26 to 133 (per Shah, Co-I Burge, and colleagues) [49–53].

#### Improvement in A1c, BMI, and PHQ-9

For our secondary outcome of successful diabetes self-management, we used A1c, BMI, and a depression scale score from the patient health questionnaire (PHQ-9) [54–56]. We chose A1c because our patients worry a great deal about their A1c levels and because it is the widely accepted standard for assessing glycemic control. To enhance the scientific quality of our research design, we added BMI as a proxy for improved diet and physical activity. Our advisors also identified depression as an important issue, so we included a depression scale score.

1. <u>Hypothesis</u>: The CCM model would result in a larger decrease in percent *A1c* from baseline to 6 months as compared to the DSMS. Previously published studies and institutional experience evaluating culturally competent diabetes management programs report changes in percent A1c with Cohen’s *f* effect sizes of 0.01 to 0.06 in studies ranging in sample size per arm from 26 to 133 [50–53].
2. <u>Hypothesis</u>: The CCM model would result in a larger decrease in BMI from baseline to 6 months than DSMS with a clinically meaningful difference of 1.5 kg/m2 between the groups (Cohen’s *f* ES = 0.06) [57,58].
3. <u>Hypothesis</u>: Compared to DSMS, CCM would result in a larger decrease (by 3 points) in PHQ-9 scores from baseline to 6 months (Cohen’s *f* ES = 0.06) [53,59,60].

## Methods

### Research setting

New Mexico, one of six majority-minority states (as of July 2019), has the largest percentage of Latinxs in the United States (46.3%). Of New Mexico’s nearly one million Latinx residents, most are of Mexican ancestry (62%) [61]. In Albuquerque, 47% of the population is Latinx. In New Mexico, 18.9% of Latinxs live in households below the FPL and 12.4% of Latinxs have diabetes.

### Participants

#### Dyadic enrollment design

We enrolled participants as dyads, where half were patients diagnosed with diabetes or pre-diabetes (“patient participants”) and half were individuals identified by the patient participant as someone in their lives who provides them with significant social support (“social support participants”). This research design was developed to respond to patient advisory board input that the importance of social relationships in Latinx culture tends to be ignored in both health research and health care. Including patient-social support dyads was a mechanism for incorporating the social dimension of patient’s lives in our research. Patient participants provided data related to their own health or experience. Social support participants primarily provided data on their perspective on their patient partner’s health.

#### Patient participants

Patient participants were individuals entering one of the two programs during the period of the study either because they were newly diagnosed with diabetes or pre/diabetes, or because they were having trouble managing their blood sugar levels. They were entering a program in order to obtain skills and information to improve their knowledge about diabetes and increase their capacity for self-management. Patient participants were adults (≥18 years old), self-≥ identified as Latinx(a), self-reported household income below 250% of the FPL, and were able to identify a social support who was willing to participate in the study. We used 250% of FPL because research has shown that individuals at 250% of the FPL are still “poor” in that they cannot afford all of the basic necessities for a healthy life [62]. Therefore, all patient participants in the study were “low-income.”

#### Social support participants

The only requirements for social support participants were that they had to be adults (18 ≥ years old) and had to be willing to participate in the research. The social support did not have to be an actual “caregiver” to the patient, nor were they screened for ethnicity or income in order to participate.

### Data collection

#### Data collectors

Three patient stakeholder data collectors [1,34,63] were trained in survey administration, the collection of biological samples (phlebotomy), and in research methods, including human subjects ethics, research protocols, consenting procedures, how to use a tablet to gather survey responses, and to use Realtime Electronic Data Capture (REDCap), the secure data capture system used and supported by the [BLINDED]. All three data collectors were native Spanish speakers. One was completely bilingual in Spanish and English, one was semi-fluent in English, and one was a monolingual Spanish speaker. Participants chose the language (English or Spanish) used for consent and data collection appointments, and data collectors were assigned accordingly.

#### Timeframe

Data were gathered at baseline between February 2016 and March 2020 [1]. Each participant was enrolled in the study for 12 months. Some 12-month data collection appointments were prematurely halted two weeks early in March 2020 due to the COVID-19 pandemic.

#### Data collection frequency

We gathered data from all participants at four time points (baseline, three months, six months, and 12 months). Baseline collection occurred before the patient participant began any program activities. All participants received a $50 merchandise card for attending each data collection appointment.

#### Data sources

We used four data sources: surveys, physical measures (A1c and BMI), patient participant and social support participant interviews/focus groups, and a program assessment of each site.

##### Surveys

At baseline, all participants were asked demographic questions. At all four time points, we asked questions from validated survey instruments.

- To measure patient understanding of what diabetes is, we used the Diabetes Knowledge Questionnaire (DKQ) [45–48].
- To measure patient ability to self-manage their diabetes, we used the Patient Activation Measure 10 (PAM-10) [38,64–71].
- To measure patient depression levels, we used the nine-question version of the Patient Health Questionnaire (PHQ-9) [54–56].
- To measure patient and social support perception of and experience with the cultural competence of their program, we used the Consumer Assessment of Healthcare *Providers and Systems Cultural Competence Set* (CAHPS-CC)] (not asked at baseline). Two additional PAM-style questions were added regarding social support.

Patient participants were asked questions about their own health. Patient participants were asked to respond to questions from their own perspective. Social support participants were asked to respond to demographic questions and the diabetes knowledge about themselves, but to answer the rest of the questions based on their perception of their patient participant partner.

##### Physical measures

At all four time points, we gathered blood samples and took height and weight measurements from patient participants only.

- A1c. The phlebotomy-trained Patient Stakeholder Data Collectors (PDCSs) drew blood samples that were tested at a UNM lab. For A1c (>10), we notified the participant.
- BMI. The PDCS documented patient participant height and weight using a standardized protocol. Height measurements were collected using SHORR boards against flat walls on level, firm (not carpeted) flooring. Weight measurements were collected using calibrated, research-grade scales. Two measurements were at each data collection point, and BMI was an average of the two measures.

##### Interviews and focus groups with participants

Data from interviews and focus groups gathered by bilingual staff were used to provide qualitative information for all three aims.

Scheduling of participants for either an interview or a focus group was based on logistical considerations related to participant availability. Interviews were conducted with individual participants; in some cases, we interviewed both the patient and social support participants. Focus groups of six to eight participants included both PPs and SPPs. Questions (Appendix) in interviews and focus groups were created to be very similar. Sessions were audio-recorded and transcribed and lasted one to two hours. Participants received a $50 merchandise card.

##### Program assessment

To assess the program interface with PP culture and context as a measure of cultural competence and “situatedness,” our program interface assessment used four sources:

- ***Inventory cataloging program components and information.*** We cataloged components of each site regarding program design, size, structure, operation, and theoretical/philosophical orientation, professional qualifications/training of program providers, activities or resources available through the programs, strategies in place for Spanish language use or acceptance, inclusion of social support participants and family, accommodation of challenges created by patient participants’ limited socioeconomic circumstances, the inclusion of stress management techniques, and data on referrals to the program, sign-ups, participation, no-shows, and attrition.
- ***Interviews with program staff.*** We conducted interviews with all relevant staff and providers to obtain their perspectives on the implementation of the programs.
- ***Patient and social support participant interviews and focus groups.*** We used data from the interviews and focus groups described above to assess participant perceptions of program interface. The questions (Appendix) included domains related to respectful treatment, language, and perspectives on the operation of the program, including the factors that were most helpful and what was missing or could be improved.
- ***Cultural competence survey*.** As indicated, the *Consumer Assessment of Healthcare Providers and Systems Cultural Competence Set* (CAHPS-CC) has questions about the participant’s experience with and perception of the program.

We made three changes to our original study protocol. One DKQ question was excluded because of lack of clarity in the wording of the question. The PAM-10 was used instead of PAM-13 to reduce burden on participants.After conducting interviews in Years 1 and 2, we reached thematic saturation and, with the approval of our PCORI Program Officer, determined not to conduct further interviews.

### Data management

The data were stored in REDCap and were assessed for quality and consistency. Analysis variables were created, including BMI categories, instrument summary scores, and income-to-federal poverty level ratios. Data were anonymized by removing demographic identifiers and randomly shifting all dates associated with a patient record within 180 days for each patient participant.

Care was taken during data collection to mitigate missing data. The largest set of missing responses came from participants unable to complete 12-month follow-up appointments due to the COVID-19 pandemic (7 of 98 at CCM from 6 to 12 months, Fig 2^1^). Missing values were imputed using the method “Multivariate Imputation by Chained Equations” via the mice R package with separate patient and social support blocks [73].

Fig 2. **Diagram of enrollments (CONSORT diagram).** DSMS, diabetes self-management support empowerment model; CCM, chronic care model.

## Qualitative Analysis

We conducted a rigorous, disciplined, empirical analysis of qualitative data using Hammersley’s criteria for qualitative research based on plausibility, credibility, and relevance [74]. We conducted a theory-driven qualitative content analysis according to standards developed by Gläser and Laudel]. Three members of the research team (two bilingual) read through transcripts to identify conceptual categories and patterns related to specified domains of inquiry, and created a qualitative codebook. We explored interconnections between theme categories and developed a holistic interpretation of the data (“constant comparison”).

## Quantitative Analysis

### Sample size and Power Calculations

Our goal was to recruit N=240 patient-social support pairs (N=120 per site) in order to obtain complete data on N=96 pairs per site, assuming a 20% attrition rate. A power analysis [63,75] showed that N=96 provided at least 80% power to detect hypothesized changes over time (described below) in the longitudinal analyses for each of the two primary endpoints (DKQ and PAM-10) and for each of the three secondary endpoints (A1c, BMI, and PHQ-9). Comparing response changes on the DKQ, PAM-10, and PHQ-9 from baseline to 6 months between the CCM to the DSMS, the two-sided Type I error rate was adjusted for the number of comparisons made (two comparisons for the co-primary outcomes) using a Bonferroni correction (α=0.025). The power analyses for detecting site differences among change scores were based on multiple linear regression models including demographic characteristics, participants’ perceived cultural competence of providers (CAHPS-CC), and social supports’ change scores on the DKQ, PAM-10, and PHQ-9 as covariates. We report Cohen’s *f* effect sizes based on the regression method, where Cohen’s standards for “small”, “medium”, and “large” effects are 0.02, 0.15, and 0.35, respectively [76,77].

The power for the primary endpoints with n=96 per site and α=0.025 for comparing the CCM to the DSMS was as follows

1. Change in DKQ summed score: Δ_CCM-DSCS_ = 2.2 (SD = 3.8), power = 96%, Cohen’s *f* effect size (ES)=0.09
2. Change in PAM-10 raw score: Δ_CCM-DSCS_ = 12.7 (SD = 24.8), power = 85%, Cohen’s *f* ES=0.07

The power for the secondary endpoints with n=96 per site and CCM to the DSMS was as follows α =0.017 for comparing the CCM to the DSMS was as follows

1. Change in A1c: Δ_CCM-DSCS_ = -0.5 (SD=1.0), power = 84%, Cohen’s *f* ES=0.06.
2. Change in BMI: Δ_CCM-DSCS_ = -1.5 (SD=3), power = 84%, Cohen’s *f* ES=0.06.
3. Change in depression scores (PHQ-9): Δ_CCM-DSCS_ = -3 (SD=6), power = 84% Cohen’s *f* ES = 0.06.

### Statistical analysis

Descriptive statistics were calculated to summarize PP characteristics. Medians and interquartile ranges (IQR) were calculated for continuous variables and were compared across sites by Kruskal-Wallis test. Frequencies and percentages were calculated for categorical variables and were compared with the chi-square test. Significant differences are noted.

Mean outcomes for Aims 1 and 2 over time and by program were analyzed using longitudinal linear mixed modeling [78,79] to account for the repeated measure effects, as well as to adjust for patient participant demographic characteristics. To adjust for potential differences in the program populations, we adjusted for potential confounding covariates identified *a priori* including patient participant gender, age, education, CAHPS CC A&B. Patient participant-Provider Communication, F. Equitable treatment, G. Trust, and H. Interpreter Services (used during a patient-provider visit); Income-to-Federal poverty level ratio, BMI, social support diabetes knowledge (DKQ-23), and social support activation (PAM-10), as well as interactions of each of these variables with time and with program. Secondary outcomes were also adjusted for patient diabetes knowledge (DKQ-23), patient activation (PAM-10), and depression (PHQ-9), except when PHQ-9 was the outcome, as well as all interactions with both time and program, since knowledge, activation, and depression can affect behavior and influence the secondary outcomes. The interaction between time and program model was of primary interest to assess whether each outcome changed to different extents over time by program. Covariates that were excluded because of their strong relationship with site included primary language and type of insurance. The models used an unstructured covariance over time. The full models were fit and then reduced with backward model selection using conditional Akaike information criterion (cAIC)] Model fit assumptions on the residuals were equal variance and normality, which were both assessed visually. However, results were robust to violations of model distributional assumption. Analyses were performed in R 4.1.0. The restricted maximum-likelihood (REML) adjusted least-squares mean difference estimate of the outcome between programs from baseline to 6 months is reported along with its 95% confidence interval. The difference estimate from baseline to 12 months is also reported.

Exploratory, post-hoc subgroup analyses summarized longitudinal changes for each outcome (PAM, A1c, PHQ-9, and BMI) by program first by each outcome’s categories, then with respect to the categories of A1c.

We conducted an analysis to assess patient participant capacity for diabetes self-management, our primary patient-reported outcome, by measuring diabetes knowledge using the DKQ and patient activation using the PAM-10 (described above). Descriptive statistics including means, standard deviations, medians, and quartiles were calculated for each outcome measure. Diabetes self-management program models were compared for the CCM *vs* the DSMS. P-values were compared to a Bonferroni-corrected α= 0.025 to account for multiple comparisons (two primary outcome measures). The primary outcome analyses were:

1. <u>Improvement in diabetes knowledge</u>: Primary analysis used the DKQ summed score.
2. <u>Improvement in diabetes-related “patient activation</u>”: Primary analysis used the PAM-10 raw score. We also converted raw scores to scaled scores per a proprietary algorithm [80] and then grouped them into patient activation levels which are displayed descriptively (frequencies and percentages) by diabetes self-management program model.

For our secondary outcome, successful patient participant management of their diabetes, we measured their A1c from blood samples drawn, PHQ-9 depression scores, and BMI calculations. The modeling used the longitudinal mixed model with the additional covariates of DKQ and PAM-10 for all three secondary outcomes and also PHQ-9 for A1c and BMI. Difference estimates are reported along with their 98.3% confidence intervals (the significance level includes a Bonferroni adjustment for three secondary outcomes).

As part of our characterization of programmatic interface with patient participant culture and socio-economic circumstances (Aim #3), we summarized patient participant and social support participant scores on five of the eight subscales of the Consumer Assessment of Healthcare Providers and Systems Cultural Competence Set (CAHPS-CC) (Domains A. and B., F., G., and H.) aggregated over the three follow-up timepoints and treated as a fixed covariate to assess the overall program experience. Medians and quartiles of the subscales by diabetes self-management program model were reported. Social support participant scores were combined with the patient participant scores and used as covariates in the analysis for the quantitative primary and secondary outcome measures.

Poverty and gender were included as covariates in the longitudinal modeling of the primary and secondary outcome variables to assess effects due to differences in poverty. Furthermore, we also assessed poverty status for potential heterogeneity of treatment effect using a separate model that included only poverty(categorical: <FPL 644 versus ≥ FPL to 250% FPL),gender, and language as covariates.

## Results

### Sample

We enrolled 452 (226 dyads of patient participants and social support participants) in the study: 120 dyads from the DSMS and 106 from the CCM). The CONSORT diagram shows the study flow from eligibility through the 12-month assessment (Fig 2). At recruitment, 58 patient participants were ineligible. The primary reasons were: no social support participant available to participate with them (n=19), already had begun the program (n=17), and income was above 250% of FPL (*i.e.,* not low-income) (n=12). We also had 84 people decline to participate. The primary reasons were: did not respond to contact call (n=28), not interested (n=18), and too busy (n=14). We enrolled more patient participants (n=120) from CDE than from One Hope (n=106), but at each site, we enrolled and retained enough patient participants to power the study (Fig 2).

Eight patient participants (3.5%) and 20 social support participants (8.8%) left the study. Attrition was not equal, with more leaving from UNM-CDE, but given the low attrition rate overall and at both sites, this was not considered a source of bias in interpreting the study findings. The primary reason for leaving was lack of time. Patient participants were eligible to stay in the study even if their social support participant partner left. However, social support participants were not allowed to stay in the study if their patient participant partner left. Included in the 20 social support participants who left were the eight whose patient participants left.

### Descriptive Baseline Statistics

#### Demographics

The majority of participants were female (72.6%) (Table 1a). Response options for the demographic question about gender were “male/female/other.” One participant identified their gender as “other.” In an interview conducted with this participant, she self-identified as a trans woman (assigned male at birth) who has not opted to initiate medication or surgical intervention for gender dysphoria. In research that is survey-based and self-report, there is movement to count trans individuals as the gender with which they self-identify, but in research dealing with biology/physiology, there is precedent for dropping them from the analysis. This project gathered both survey and biological data. We made a conscious decision not to exclude this one individual from the analysis, as that would align with the binary gender tradition in research that reflects and contributes to social and political dynamics that make transgender individuals invisible. Because we would like to honor this participant’s gender, as she states, we present gender as a non-binary variable both out of respect and to represent her in the research as she represents herself. However, for statistical purposes, rather than dropping her from the analysis, she was coded as “male.” We made this decision because she was diagnosed with diabetes long before publicly self-identifying as a trans woman and she has not made any physiological changes.

**Table 1a.**
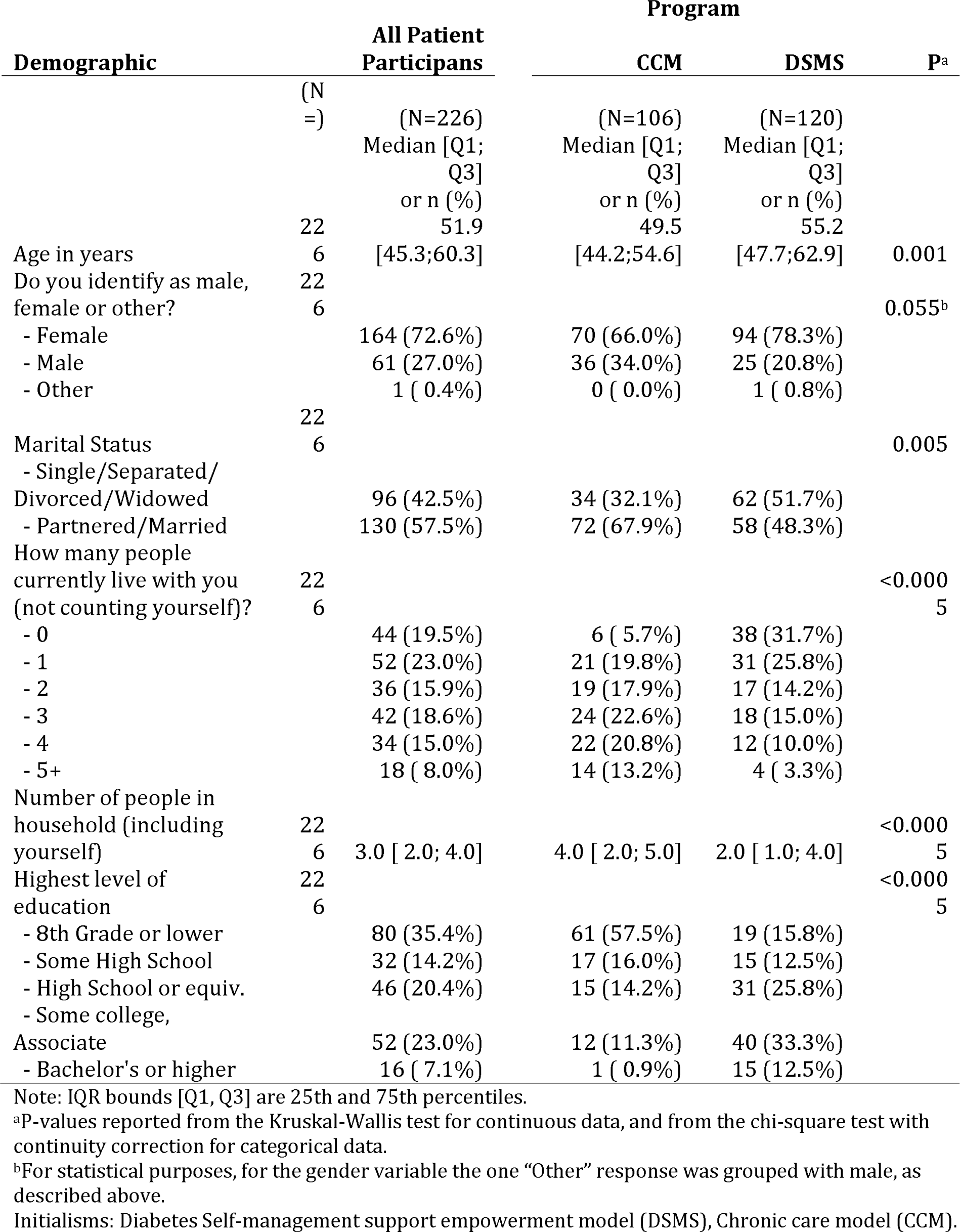
Baseline patient participant demographics (continued in Table 1b).

Therefore, we hypothesized that her diabetes-related biology would be more in alignment with male physiology than female. The majority of participants were married (57.5%) and reported speaking Spanish as the only language at home (58.4%) (Table 1a). The median age of the patient participants was 51.9 (IQR bounds: 45.3 to 60.3) years. The household size was fairly evenly distributed among living alone or having one to four other household members (range: 15%-23%). Nearly one-half (49.6%) did not graduate from high school. The majority of participants self-rated their health as either “*good*” (44.4%) or “*fair*” (32.9%) (Table 1b).

**Table 1b.**
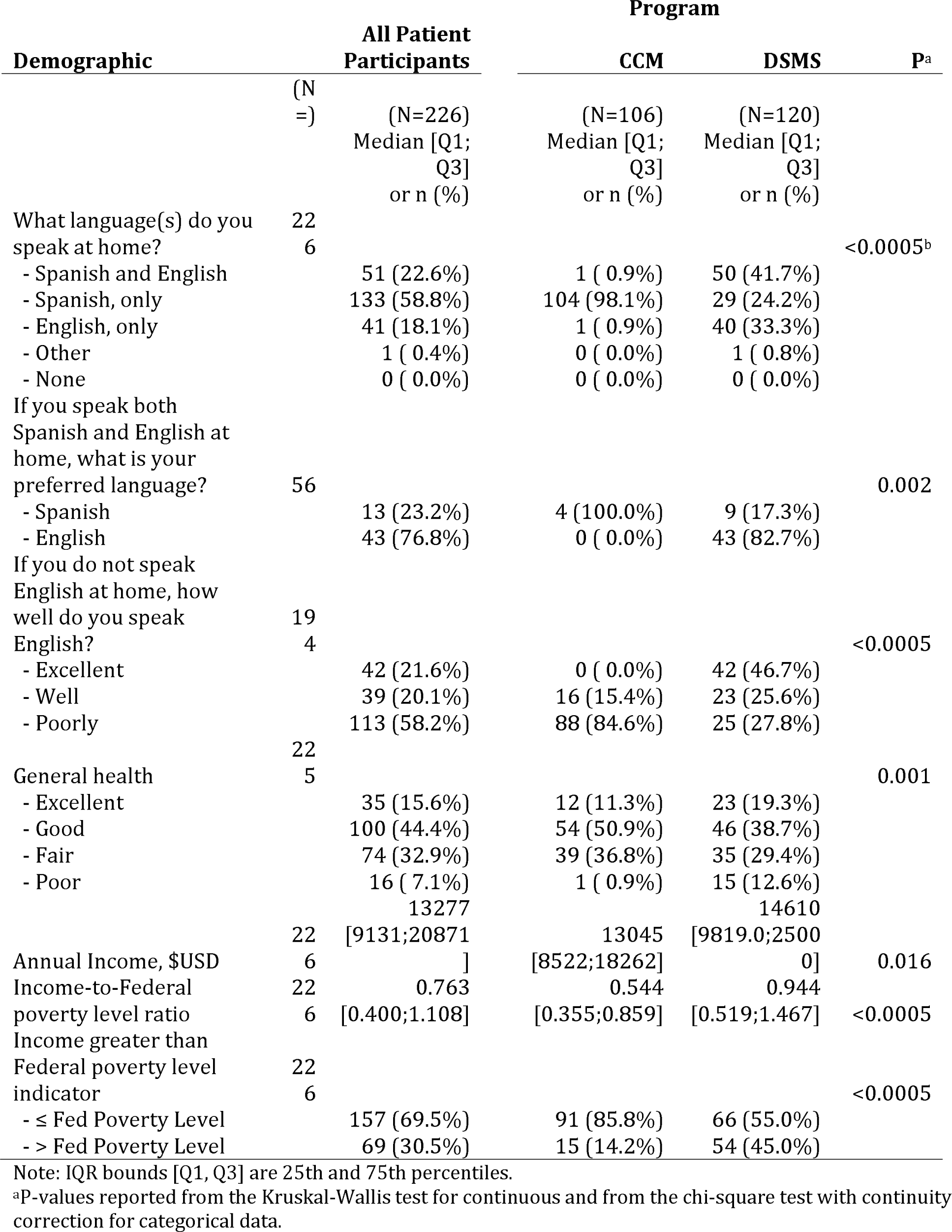
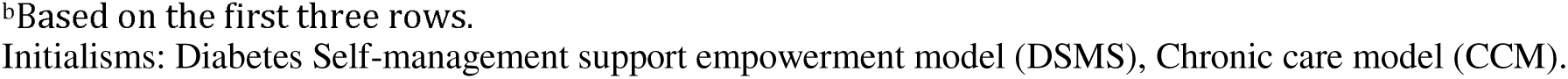
Baseline patient participant demographics (continued from Table 1a).

Compared to patients in the DSMS, patients in the CCM were significantly younger (49.5 vs 55.2), slightly more likely to be male (34% vs 21%), more likely to be partnered or married (67.9% vs 48.3%), and less likely to have graduated from high school (73.5% vs 28.3%) (Table 1a). CCM patients were also significantly more likely to only speak Spanish at home (97.2% vs 24.2%), less likely to prefer to speak English if they speak both Spanish and English at home (0% vs 82.7%), and less likely to speak English well if they do not speak English at home (15.4% vs 72.3%). CCM patients had much lower income-to-FPL ratios (0.54 vs 0.94) and were more likely to live no higher than FPL (85.8% vs 55.0%) (Table 1b).

#### Physiological characteristics

Patient participants were split between those who entered with pre-diabetes (42.2%) and diabetes (57.8%), with the median time since diagnosis of their current diabetes category 1.6 years (Table 2a). Some patient participants had recently tested in the pre-diabetes range which was why their provider had referred them to a program but between diagnosis and when they began the program, some made lifestyle changes such that when we drew blood samples for A1c at baseline, they were below the range for prediabetes. We did not exclude them from the study because the A1c analysis conducted as part of our baseline because we were not using an A1c test to screen for enrollment.

**Table 2a.** Baseline diabetes, BMI, Diabetes knowledge, and patient activation characteristics (continued in Table 2b).

| Diabetes | (N=) | All Patient<br>Participants<br>(N=226)<br>Median [Q1;<br>Q3]<br>or n (%) | Program |  | P<sup>a</sup> |
| --- | --- | --- | --- | --- | --- |
|  |  |  | CCM<br>(N=106)<br>Median [Q1;<br>Q3]<br>or n (%) | DSMS<br>(N=120)<br>Median [Q1;<br>Q3]<br>or n (%) |  |
| Diabetes diagnosis | 218 |  |  |  | 0.011 |
| - Pre-diabetes |  | 92 (42.2%) | 52 (52.0%) | 40 (33.9%) |  |
| - Diabetes |  | 126 (57.8%) | 48 (48.0%) | 78 (66.1%) |  |
| Time since diagnosis (years) | 214 | 1.55 [0.08;8.01] | 1.00 [0.08;4.00] | 2.00 [0.08;12.01] | 0.028 |
| Baseline A1c | 211 | 6.6 [ 6.0; 9.3] | 6.5 [ 5.9; 8.8] | 6.7 [ 6.0; 9.4] | 0.347 |
| A1c Categories | 211 |  |  |  | 0.636 |
| - Neither |  | 14 ( 6.6%) | 8 ( 7.5%) | 6 ( 5.7%) |  |
| - Pre-diabetes |  | 77 (36.5%) | 41 (38.7%) | 36 (34.3%) |  |
| - Diabetes |  | 120 (56.9%) | 57 (53.8%) | 63 (60.0%) |  |
| Body mass index (BMI) | 226 | 32.7 [29.2;36.9] | 31.3 [28.2;36.4] | 33.8 [30.1;37.1] | 0.036 |
| Body mass index (BMI)<br>Categories | 226 |  |  |  | 0.041 |
| - Underweight |  | 2 ( 0.9%) | 0 ( 0.0%) | 2 ( 1.7%) |  |
| - Normal or Healthy Weight |  | 21 ( 9.3%) | 11 (10.4%) | 10 ( 8.3%) |  |
| - Overweight |  | 48 (21.2%) | 30 (28.3%) | 18 (15.0%) |  |
| - Obese |  | 155 (68.6%) | 65 (61.3%) | 90 (75.0%) |  |
| Are you currently taking any<br>medications for diabetes, any<br>of the conditions you just<br>mentioned, or for anything<br>else?<sup>b</sup> | 213 |  |  |  | <0.0005 |
| - Yes |  | 160 (75.1%) | 61 (62.9%) | 99 (85.3%) |  |
| - No |  | 53 (24.9%) | 36 (37.1%) | 17 (14.7%) |  |
| Are you taking your<br>medication(s) as prescribed? | 156 |  |  |  | 0.031 |
| - Yes |  | 132 (84.6%) | 56 (93.3%) | 76 (79.2%) |  |
| - No |  | 24 (15.4%) | 4 ( 6.7%) | 20 (20.8%) |  |
| Have your (biological) family<br>members been diagnosed<br>with diabetes? | 223 |  |  |  | 0.301 |
| - Yes |  | 182 (81.6%) | 90 (84.9%) | 92 (78.6%) |  |
| - No |  | 41 (18.4%) | 16 (15.1%) | 25 (21.4%) |  |

Note: IQR bounds [Q1, Q3] are 25th and 75th percentiles.
<sup>a</sup>P-values reported from the Kruskal-Wallis test for continuous and from the chi-square test with continuity correction for categorical data.
<sup>b</sup>Medications were not reported separately for diabetes versus other conditions.
Initialisms: Diabetes Self-management support empowerment model (DSMS), Chronic care model (CCM).

Median baseline A1c was 6.6, with 33.3% of those with diabetes having A1c over 10. Median BMI was 32.7 (Table 2a). The vast majority reported a family member with diabetes (81.6%), were taking some type of medication (75.1%), and that their social support participant partner was helpful to them in managing their diabetes (90.7%).

Patient participants at both sites had comparable median A1c values (CCM 6.5% vs DSMS 6.7%) with a similar distribution over A1c diabetes ranges of pre-diabetes and diabetes, and similar median BMI (31.2 vs 33.6) (Table 2a). The majority in both programs were above normal BMI compared to norms for American adults--between 18.5 and 24.9. For CCM, 61% and for DSMS, 90% were obese. There were differences between PPs in the CCM and those in the DSMS on variables such as entering the program with diabetes, time since diagnosis, taking medications for diabetes, adhering to the medication prescription, and the importance of the social support in managing the disease.

#### Capacity for diabetes self-management

All patient participants generally scored low on diabetes knowledge (DKQ), with an average score of 14/23, and compared to DSMS, patient participants in CCM scored lower by two points (13 vs 15) (Table 2b). However, these values were consistent with baseline assessments of similar populations.

**Table 2b.**
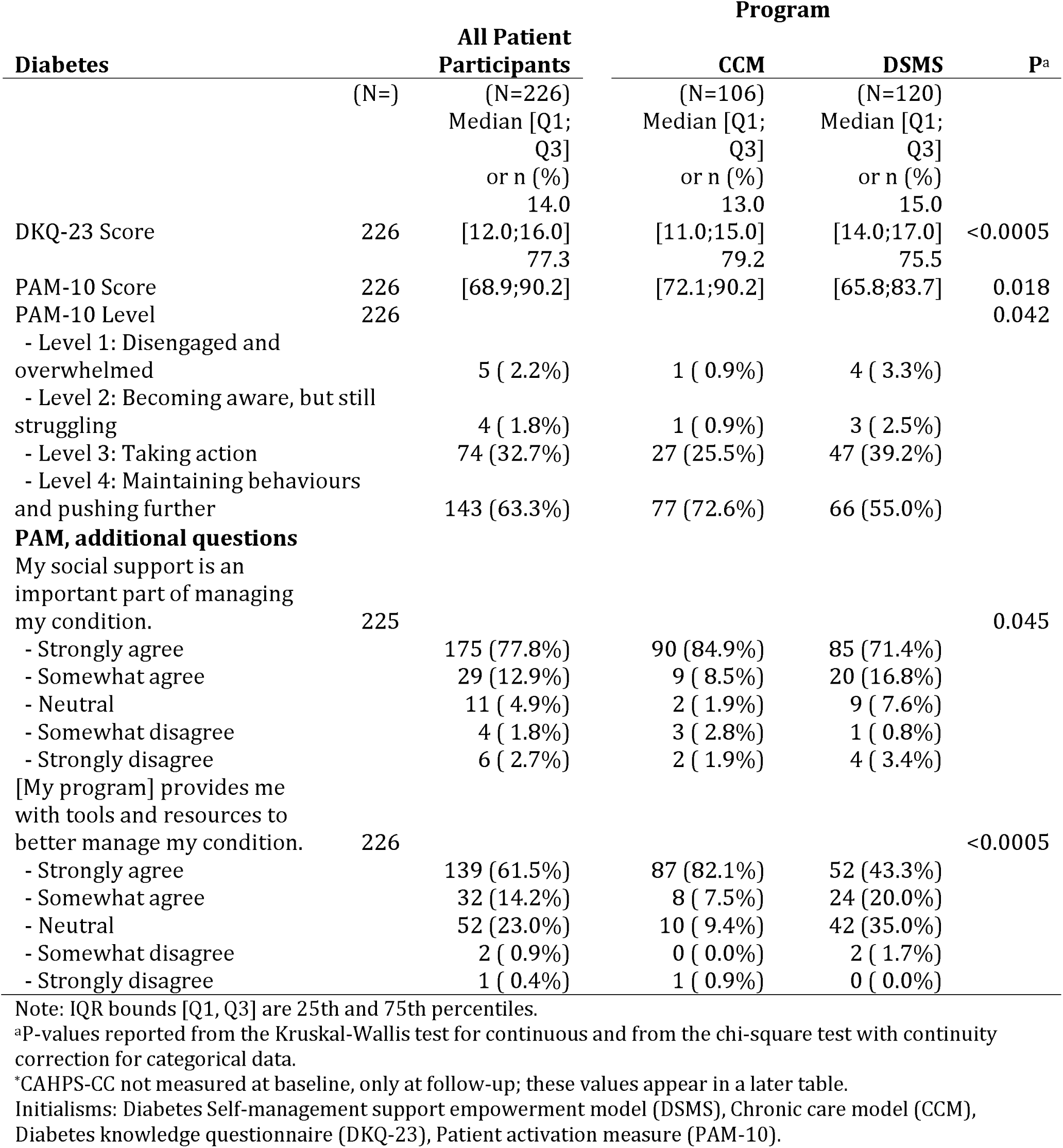
Baseline diabetes, BMI, Diabetes knowledge, and patient activation characteristics (continued from Table 2a).

The average PAM-10 score was 77.3, with 96% scoring at Level 3 (taking action) or 4 (maintaining behaviors/pushing further) (Table 2b). PPs in CCM compared to patient participants in DSMS scored higher by four points with more patient participants at Level 4 (72.6% vs 55.0%) and with fewer at Level 1 (disengaged/overwhelmed) and Level 2 (becoming aware, but struggling) (1.8% vs 5.8%).

#### Depression

More than half (52.2%) of the patient participants ranked as having symptoms of depression, with 22.2% in the “Moderate,” “Moderately Severe,” or “Severe” categories (Table 3). More than half in either program scored as having mild to severe depression although a higher percentage of subjects in DSMS tested for moderately severe to severe depression (11.7%) compared to CCM (3.8%).

**Table 3.**
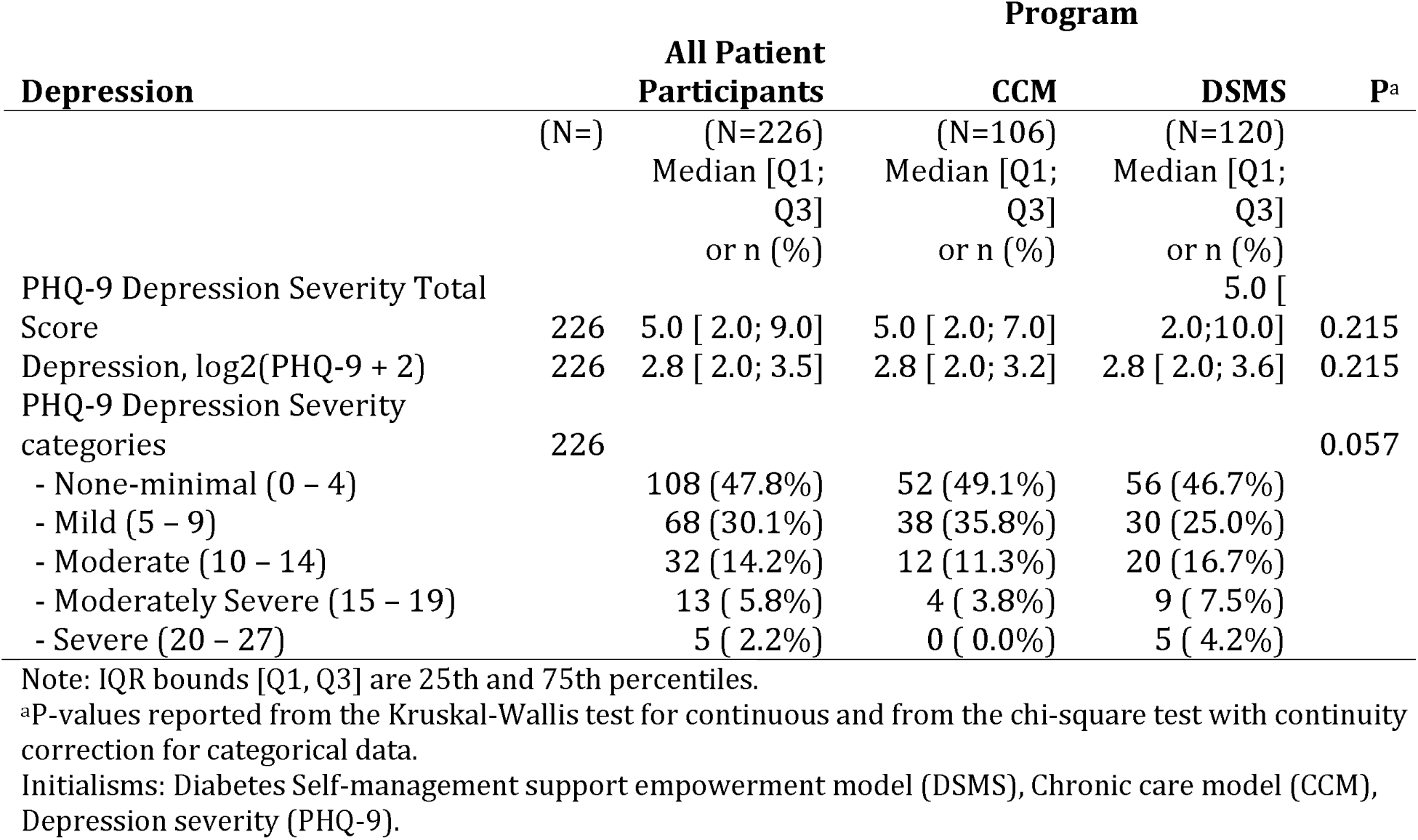
Baseline depression characteristics.

#### Selected environmental and comorbid conditions

The three most commonly reported comorbid conditions reported were hypertension (28.3%), high cholesterol (16.4%), and a thyroid condition (13.7%) (Table 4). Rates for hypertension between patient participants in the two programs were similar; however, CCM patient participants were less likely to have any comorbid condition (41.5% vs. 70.0%) and had fewer comorbid conditions reported (Q1-Q3: 0-1 vs. 0-3).

**Table 4.**
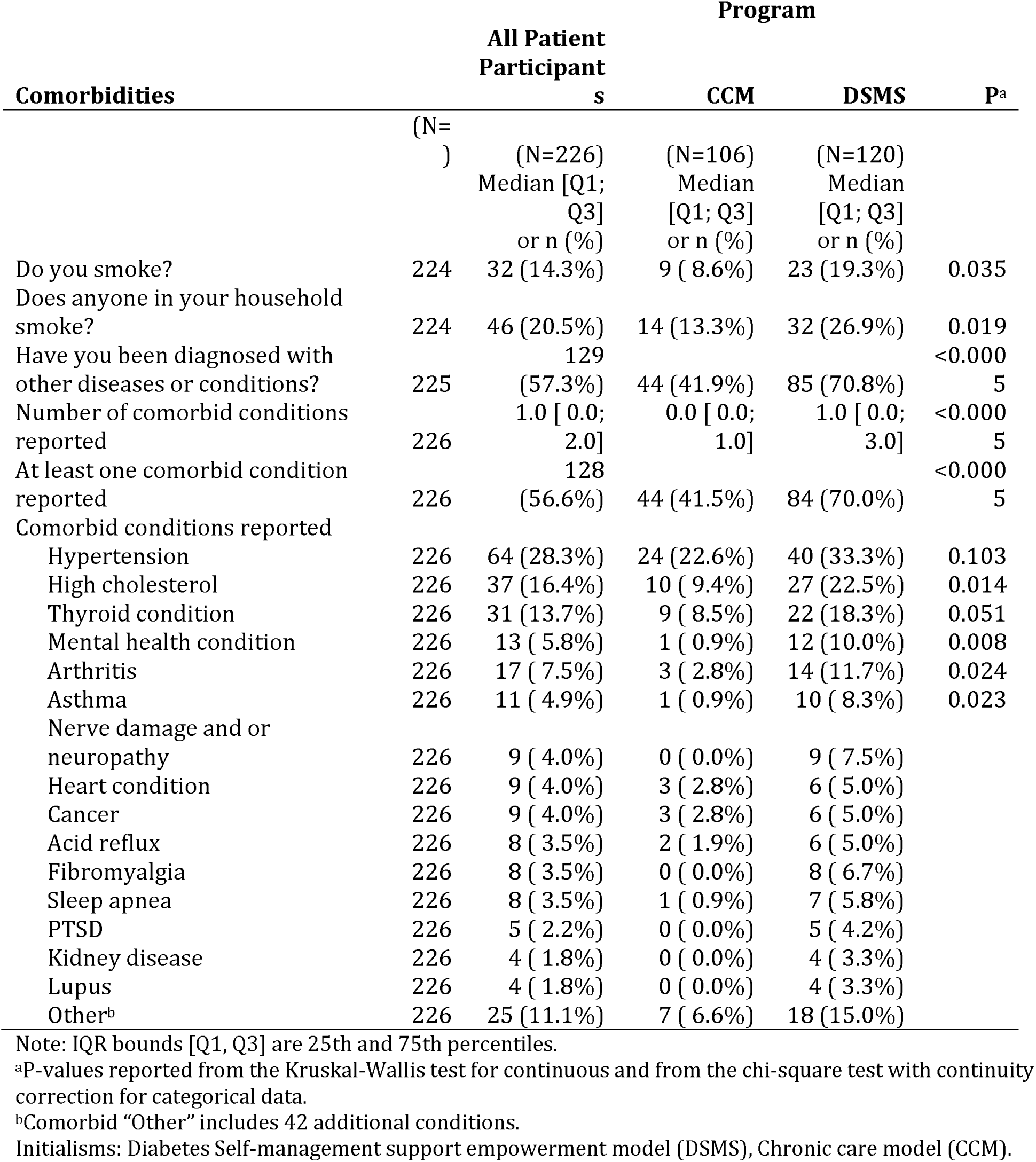
Environmental and Comorbid selected characteristics. Rare comorbid conditions excluded from table.

#### Social supports

The most common social support participant relationship categories were family member other than spouse (42.5%) and spouse (27.9%) (Table 5a). Most patient participants reported daily interaction (65.9%) with the social support participants, primarily in-person (85.8%), although only slightly more than half live in the same household with their social support participant (Table 5b).

**Table 5a.** Patient Participants’ report on social support participant characteristics (continued in Table 5b).

| Patient Participants About Their<br>Social Support Participant | (N=) | All Patient<br>Participants<br>(N=226)<br>n (%) | Program |  | P<sup>a</sup> |
| --- | --- | --- | --- | --- | --- |
|  |  |  | CCM<br>(N=106)<br>n (%) | DSMS<br>(N=120)<br>n (%) |  |
| What is your relationship to your<br>social support partner in this<br>research project? | 226 |  |  |  | 0.313 |
| - spouse |  | 63 (27.9%) | 33 (31.1%) | 30 (25.0%) |  |
| - cohabitating partner/long-term<br>partner |  | 21 ( 9.3%) | 8 ( 7.5%) | 13 (10.8%) |  |
| - family member |  | 96 (42.5%) | 39 (36.8%) | 57 (47.5%) |  |
| - friend |  | 40 (17.7%) | 23 (21.7%) | 17 (14.2%) |  |
| - neighbor or other |  | 6 ( 2.7%) | 3 ( 2.8%) | 3 ( 2.5%) |  |
| How involved is your social support<br>partner in your diabetes<br>management? | 226 |  |  |  | 0.586 |
| - very |  | 134 (59.3%) | 67 (63.2%) | 67 (55.8%) |  |
| - somewhat |  | 61 (27.0%) | 25 (23.6%) | 36 (30.0%) |  |
| - not very |  | 24 (10.6%) | 10 ( 9.4%) | 14 (11.7%) |  |
| - not at all |  | 7 ( 3.1%) | 4 ( 3.8%) | 3 ( 2.5%) |  |
| How often does your social support<br>partner accompany you to<br>appointments with your medical<br>provider? | 226 |  |  |  | 0.418 |
| - always |  | 59 (26.1%) | 30 (28.3%) | 29 (24.2%) |  |
| - often |  | 24 (10.6%) | 12 (11.3%) | 12 (10.0%) |  |
| - sometimes |  | 73 (32.3%) | 37 (34.9%) | 36 (30.0%) |  |
| - never |  | 70 (31.0%) | 27 (25.5%) | 43 (35.8%) |  |
| How much does your social support<br>partner know about your health? | 226 |  |  |  | 0.634 |
| - a lot |  | 151 (66.8%) | 68 (64.2%) | 83 (69.2%) |  |
| - some |  | 59 (26.1%) | 29 (27.4%) | 30 (25.0%) |  |
| - not much |  | 15 ( 6.6%) | 8 ( 7.5%) | 7 ( 5.8%) |  |
| - nothing |  | 1 ( 0.4%) | 1 ( 0.9%) | 0 ( 0.0%) |  |

Note: IQR bounds [Q1, Q3] are 25th and 75th percentiles.
<sup>a</sup>P-values reported from the Kruskal-Wallis test for continuous and from the chi-square test with continuity correction for categorical data.
Initialisms: Diabetes Self-management support empowerment model (DSMS), Chronic care model (CCM).

**Table 5b.**
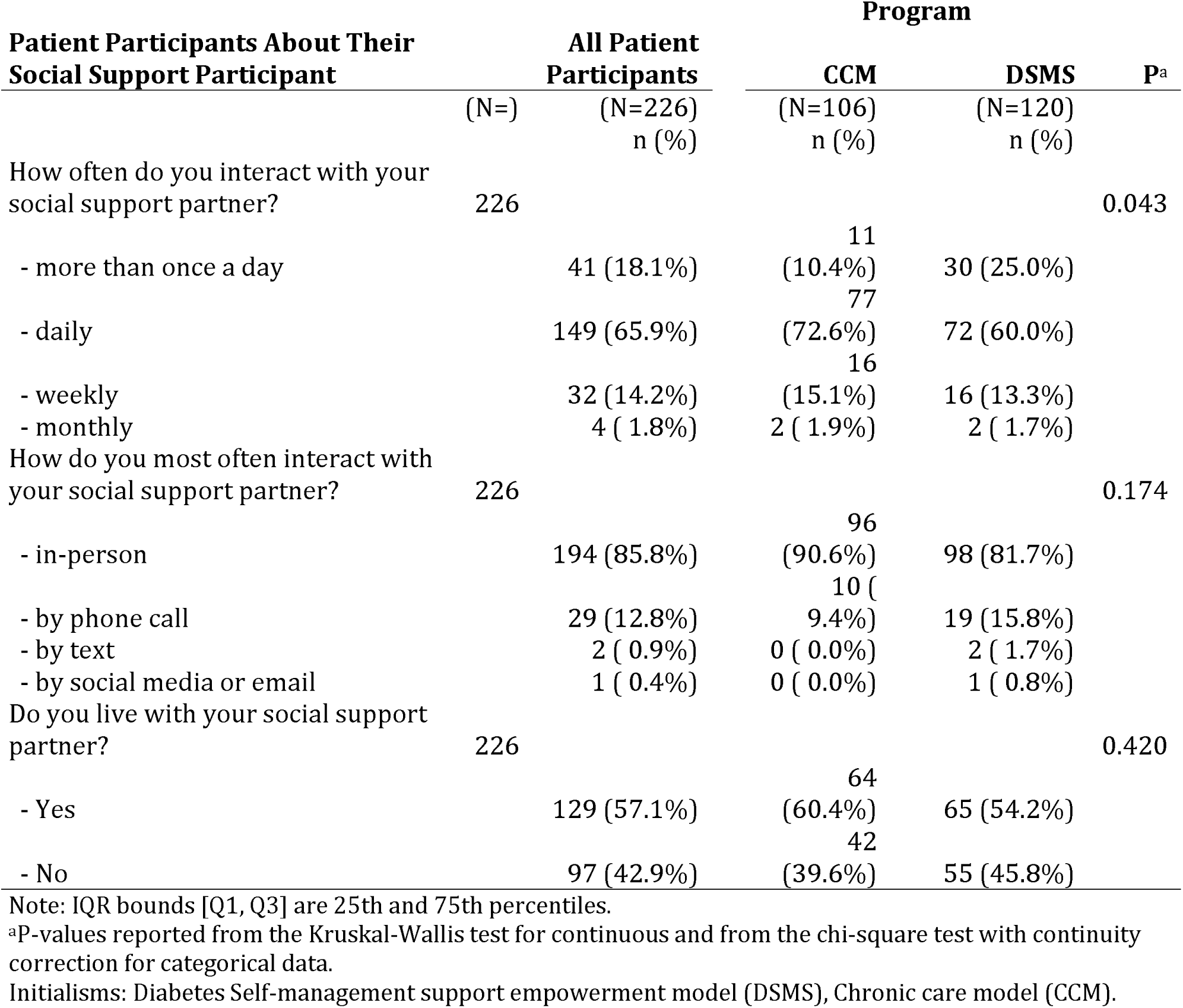
Patient participants report on social support participant characteristics (continued from Table 5a).

The mean age for social support participants was 46.3 years and the majority were female (64.6%) (Table 6). Educational attainment for social support participants was diverse, with nearly one-third having completed 8^th^ grade or lower, and nearly one-third having completed high school or the equivalent (28.8%). Eighteen social supports (8.0%) had a bachelor’s degree or higher. CCM social support participants were slightly more likely to be female (68.9% vs 60.8%). Educational attainment at the sites was somewhat different--nearly half of CCM social support participants (45.3%) had only completed 8^th^ grade or lower verses 16.7% for DSMS, and CCM social support participants were less likely to have completed high school or the equivalent (22.6% vs 34.2%), to have some college or an associate’s degree (15.1% vs 23.3%), or to have a bachelor’s degree or higher (1.9% vs 13.3%).

**Table 6.**
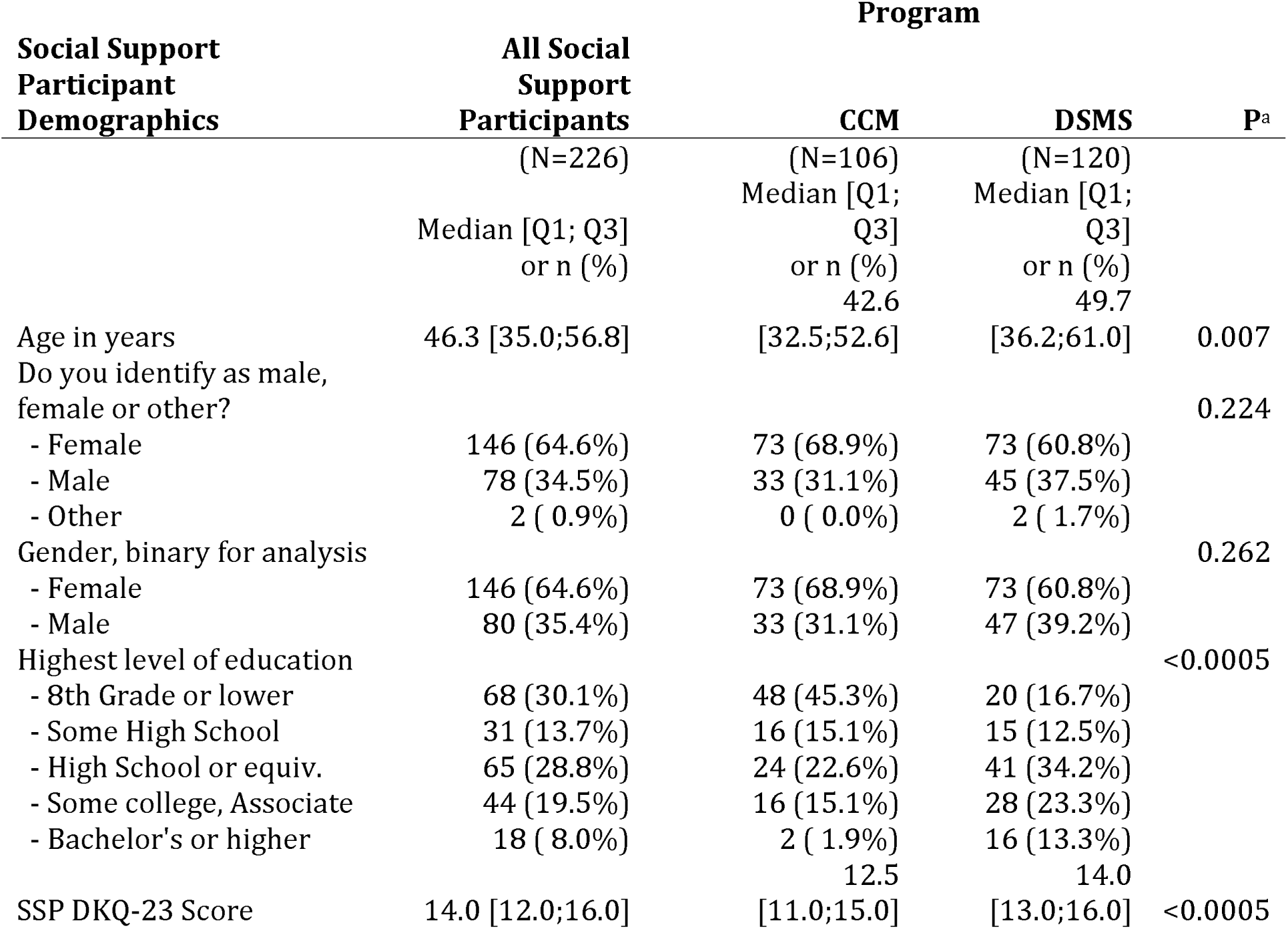
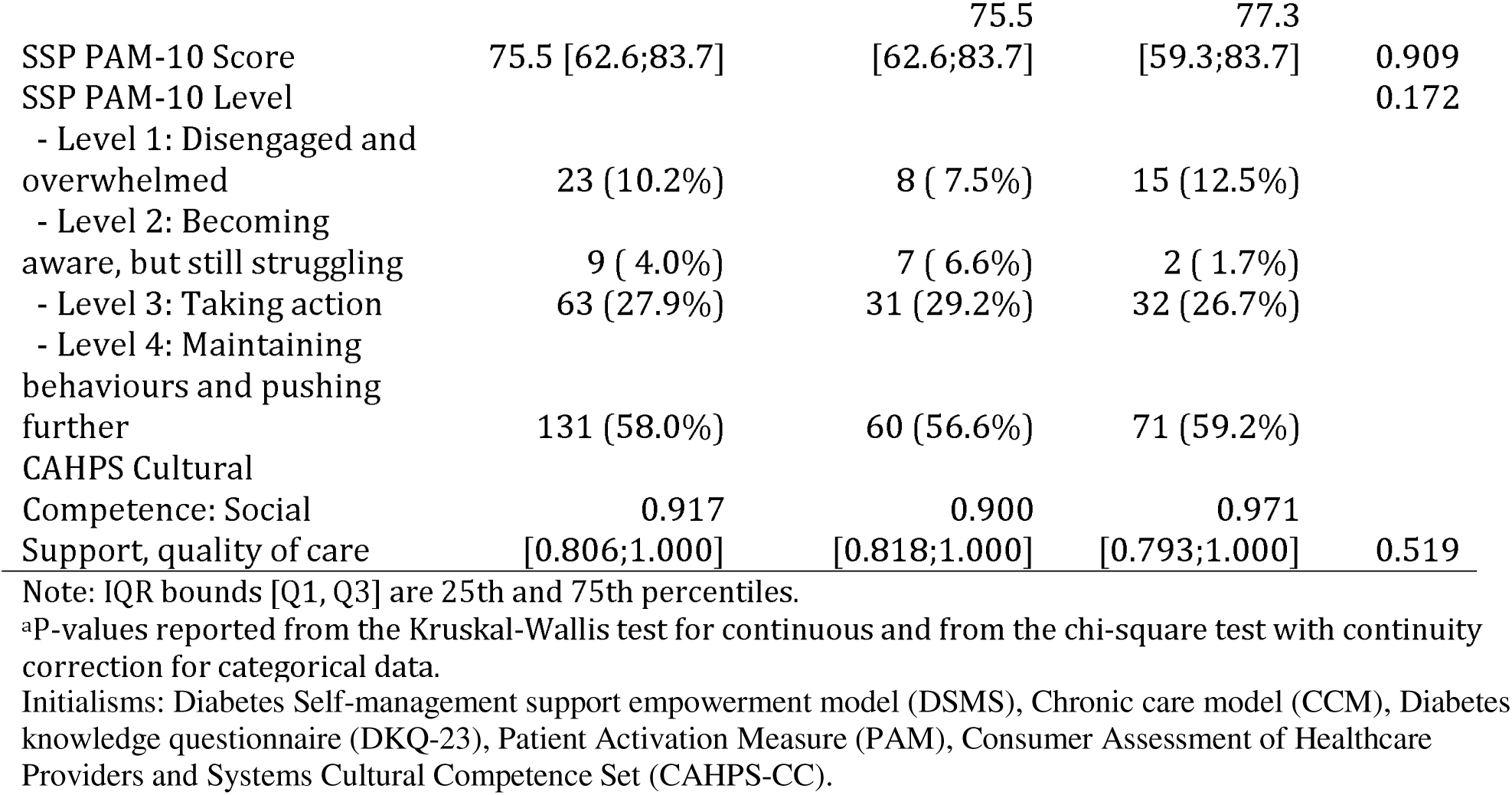
Social support demographic characteristics, diabetes knowledge scores, patient activation, and overall *Consumer Assessment of Healthcare Providers and Systems Cultural Competence Set* (CAHPS-CC) score.

### Qualitative Analysis

#### Coding of interviews and focus groups

During Year 1, we conducted 11 provider interviews—five at CCM, six at CDE. During Years 1 and 2, we conducted eight focus groups (2/site/year) with a mix of 36 patient participants and social support participants, and 44 interviews with 23 unique patient participants (12 CCM, 11 CDE) and 21 unique social support participants (12 CCM, 9 CDE). In Year 1, we identified 42 Level I qualitative coding themes. After reviewing the themes and discussion, we selected themes that were well represented and fit with our line of inquiry. We used the themes to conduct Level II coding, rereading the transcripts and extracting quotations related to each of the six themes. We cleaned each quote, removing extraneous utterances, providing parenthetical contextual cues when appropriate, and ensured that the quotes were thematically relevant and broadly represented for both sites. For Level III coding, we united extractions created by all three team members and when appropriate, we sorted them into subthemes in order to achieve better internal thematic coherence.

For Aim #1, participant interviews and focus groups informed us about participant knowledge, activation, and motivation, and the barriers to both. For Aim #2, we learned about challenges to reducing A1c, making lifestyle changes (including eating—which impacts BMI), and mental health challenges related to reducing stress and depression. For Aim #3, from program staff interviews, we learned about the specifics of each program as discussed in the program assessment section below. From participant interviews and focus groups, we learned about the importance of social support and participant attitudes toward and experience with their program.

#### Importance of social support

Participants at both sites discussed the importance of social support in helping them deal with and manage their diabetes. Support was indicated to relate to having someone who was interested in their health and well-being, and “being there” for them was identified as an important motivator. Support related to food involved helping the patient participant shop for and prepare diabetes-appropriate food, helping them develop better food-related habits (such as not keeping chocolate or junk food in the house), monitoring what the patient participant was eating or purchasing, helping them make rules or a plan for shopping and eating, scolding them if they tried to break the “rules,” and having them watch educational programs about diabetes, about the politics of the food system that contribute to disease, or about how bad junk food is for you. One key thing that many social supports do is to change their own diet and eating habits in order to support the patient participant in their struggle with diabetes. In addition, social support provided by the programs was identified as very important.

#### Motivations for change

We identified things that motivate people to lifestyle changes (Table 7), including the importance of being able to work, fear of illness or diabetes complications, and for the benefit of others, especially children. While participants at both sites shared sentiments related to fear of being unable to work, we noted a difference between the sites, with the participants at CCM expressing more concern about what might happen if they were unable to work.

**Table 7.** Motivations for change.

|  |  |
| --- | --- |
| Importance of Work | “I am so afraid that if I stop working, I’ll become crippled.” |
|  | “Diabetes as an unexpected expense, things that you didn’t expect to spend on, but it’s for our own good” |
|  | You can’t work, you don’t have a life, you don’t have anything.” |
|  | “They don’t want to take pills. They don’t want to go to a doctor because it’s too expensive. They don’t have a job. I have the desire to do it and I am interested in having a healthy life. I want to live healthy for a long time and my life is work.” |
| Fear of Illness/Symptoms | **“Sometimes I won’t finish my soda. I’ll just drink half of it and throw half of it away, because I’m thinking of the result if I did drink all that. I don’t want to get into a diabetic coma or get into any other medical issues.”** |
|  | **“...it was serious, because this time, it took some body parts. We know that we have to make the transition. We have no choice... they already got rid of all my toes...it’s going to have to come to the knee. Then, where does it stop?”** |
| For the benefit of others, especially children | “I was concerned because I said maybe I won’t take care and they’ll cut my feet. That’s what am afraid of because if you don’t take care of yourself they can cut off your feet and that’s what I don’t want. How I am going to support myself. I don’t want to ask my children because they barely have for their own things, their water, their electricity and the young people nowadays want name brand clothing and no, |
|  | no, no. I tell my children, don't give me anything, I don't need it. Because they barely have for themselves. I worried about that because I think how I am going to go to work. I have a lot of years and I worry and I say I need to do something." |
|  | "It's very scary. You know why? Because I have babies, nine grandbabies. I always say, "If God is going to give me a long life, I would like to be healthy. I don't want to be a burden on my kids or my grandkids. I want to help them. Yeah, so I'm really striving to get off the pills. In order to do that, I need to bring down my A1c. It was very high." |
|  | "[If I] just make a change so that the grandkids, they won't notice that you changed their eating habits, so that it will be easier for them as they get older. Maybe that will help control their weight and hopefully lessen their [risk of getting diabetes]." |
|  | **"Well, if I die, I'll be with the Lord, but that's selfish thinking, because I have family and they're the ones that keep me going, my grandkids and my boys. I just love them so much."** |
|  | "Since I was detected with pre-diabetes, I am watching what I eat, but everyone in the family, my children and all of us are eating the same...my oldest son said, "wow, we all have diabetes now." |

#### Understanding of/knowledge about diabetes

We identified themes related to diabetes knowledge (Table 8). Some participants had knowledge from personal experience with a family member. Many participants discussed having a lack of knowledge or poor understanding of diabetes.

**Table 8.** Diabetes Knowledge as a Theme.

|  |
| --- |
| "To be honest I don't know how to deal with this. I don't know what are the symptoms. I don't know how it affects my health. I don't have a pattern to follow." |
| "It's the ignorance, am sure everyone here knows people who are diabetics, but we don't know why how it affects and what are the consequences." |
| "That is something like I don't know how many carbohydrates, the chicken has protein. Honestly, I eat because I need to eat but not because I know how to balance or because it has these many minerals or this many proteins. To be honest I don't know anything." |
| "You are supposed to know what you eat. How am I going to know this? I just know that am hungry and I need to eat." |

#### Denial

Denial was another common theme (Table 9). Participants described how they did not take the diagnosis seriously, or felt they could not deal with the required changes and chose to ignore it or disbelieve it.

**Table 9.** Denial as a Theme.

|  |
| --- |
| “Up to this day, he (my husband) claims that he doesn’t have it...” |
| “Me, I don’t feel that I have diabetes. I don’t feel it... My life was all about work my life has been working daily, I think the body gets tired and sometimes wants vitamins and I don’t what else. That’s all I could tell you...that’s what they tell me...to be honest I feel that the cholesterol also makes you dizzy, right? You get dizzy with cholesterol...When I walk a lot, I get home tired, and dizzy but I take some pills that I got here and it goes away. I think is age related you’re old and get tired...I gain weight just sleeping and that’s where I developed it.” |
| “He doesn’t accept it, he says, I don’t have it yet, am in the borderline because when we first started coming, he was on the borderline. He was told is your decision if you want to cross the line. ...Even now he still says that he’s still on the borderline but it’s a lie because he already has it.” |

#### Barriers to change

We asked participants if it was difficult to make the necessary behavior changes and they identified three types of challenges: 1.) Exercise is hard—lack of time or energy primarily because of long work hours. 2.) Dietary changes are hard—it is “boring,” the small portions, “addiction to food,” taste, custom. 3.) Social challenges—other people don’t understand, you don’t want to offend other people, being around other people who are eating unhealthy foods makes it harder for you to avoid them. Although it can be difficult to make lifestyle changes, people see that changing habits, andeating healthier in particular, as an investment. It is a little more expensive now, but they understand that it is preventative.

#### Agency

Some participants were very motivated and felt they have a lot of control over their health. This involved acceptance and feeling they have agency to manage the condition and prevent complications if they take personal responsibility. Others described frustration or resignation—a feeling that diabetes is controlling them or that it is something external happening to them.

### Quantitative Analysis

#### Aim #1: Primary outcomes

The primary analysis for Aim #1 was to measure and compare improvement in patient participant capacity for diabetes self-management as indicated by improvement at 6 months in diabetes knowledge and patient activation. For each outcome variable the resulting model is interpreted for model fit, time-by-program interaction, main effects not associated with interactions, and interactions (Fig 3A). Model fit assumptions were not violated.

Fig 3. **Comparison of results from the Diabetes knowledge questionnaire (DKQ).** Scores are taubluated and primary contrast over time between programs. (A) Longitudinal plot. (top) The diamonds are means by program over time, thin lines are trajectories for each PP. (bottom) Marginal histograms of DKQ scores for each program and time point. (B) Black dots are least-squares means, 95% confidence intervals represented with blue bars and red comparison arrows for pairwise comparisons between means. If an arrow from one mean overlaps an arrow from another group, the difference is not statistically significant. (C) Primary contrast testing whether the change between time points (e.g., 6 mo vs 0 mo) for CCM was different than the same change for DSMS. If the 95% confidence interval does not overlap 0, then there is an (uncorrected) difference between programs over time. Since the 95% CI crosses zero in all three contrasts, there is no difference between the programs for DKQ. DSMS, diabetes self-management support empowerment model; CCM, chronic care model.

#### Changes in diabetes knowledge

Scores on the PP DKQ-23 were used to measure diabetes knowledge. Scores increased for patient participants in both programs. Both sites increased their knowledge from baseline to 3 months (approximately 0.75 to 1 point) and sustained their improvement through the end of the study. There was no Time-by-Program interaction (Fig 3C, p-value = 0.265), so both programs changed to similar extents over time; in particular, the Baseline-to-6-month change for CCM DKQ-23 (a 23-point scale) compared to the change for DSMS was 0.483, 95% CI (-0.119, 1.086). DSMS scored approximately 1.25 points higher than CCM (Fig 3B, p-value = 0.0009).

The multivariate analysis (Table 10, contrasts not shown) revealed that the PP DKQ-23 Score was positively related to SSP DKQ-23 Score (regression coefficient (coef) = 0.198, 95% CI: (0.137, 0.258)), negatively related to Age (coef = -0.0266, 95% CI: (-0.0544, 0.00127)), and positively related to Education (High School and lower categories had lower scores than Some College and above, about 1.75 difference). PP DKQ-23 Score was negatively related to the Consumer Assessment of Healthcare Providers and Systems Cultural Competence Set (CAHPS-CC) A and B Communication for CCM (coef = -0.626, 95% CI: (-1.11, -0.14)) but positively related for DSMS (coef = 0.402, 95% CI: (0.0334, 0.77)) (Fig 3).

**Table 10.** DKQ-23 outcome Type III ANOVA Table for reduced longitudinal model.

| Variables | NumDF | Pr(>F) |
| --- | --- | --- |
| Time | 3 | <0.0005 |
| Program | 1 | 0.044 |
| DKQ-23 Score, SS | 1 | <0.0005 |
| CAHPS A&B Communication | 1 | 0.469 |
| Age | 1 | 0.061 |
| Highest level of education | 4 | <0.0005 |
| Program : CAHPS A&B Communication | 1 | 0.001 |
| Time by Program | 3 | 0.265 |

Initialisms: Social support (SS), Consumer Assessment of Healthcare Providers and Systems Cultural Competence Set (CAHPS-CC) (CAHPS-CC), Diabetes knowledge questionnaire (DKQ-23).
A Type-3 test assesses whether a term explains substantial variance in the response after adjusting for the other covariates. The two primary values to interpret are the numerator degrees-of-freedom and the associated p-value for the test.

#### Patient activation

We used the PAM-10 as a proxy for measuring the patient’s capacity for self-managing their diabetes, conceptualized as patient activation. The average PAM-10 score was 77.3, with the vast majority scoring at Level 3 (taking action) or 4 (maintaining behaviors and pushing further). Patients in the CCM compared to patients in the DSMS scored higher by four points with more patients at Level 4 (72.6% vs 55.0%) and with fewer at Levels 1 and 2 (1.8% vs 5.8%) (Table 2).

We used the PAM-10 numeric score because it is more sensitive to detecting differences than rounding the scores into four activation categories. PAM-10 scores increased for PPs in both programs (Fig 4A). Model fit assumptions were not violated. There was no Time-by-Program interaction (Fig 4C, p-value = 0.612), so both programs changed to similar extents over time; in particular, the Baseline-to-6-month change for CCM compared to the change for DSMS was -1.278, 95% CI (-4.56, 2.01). DSMS PAM scores (scale from 0 to 100) were close to 77 at baseline and CCM was close to 78 at baseline, but both increased to 971 about 82 at 3 months and beyond (Fig 4B). Both sites increased their activation scores by 6 months (p-value < 0.0001). The multivariate analysis (Table 11, contrasts not shown) revealed that the Patient Activation (PAM-10) was positively related to age (0.162, 95% CI: 0.046, 0.277), negatively related to education (Bachelor’s or higher was lowest at 75, some high school through some college were medium at 80, and 8th grade or lower was highest at 86), negatively related to BMI (coef = -0.183, 95% CI: -0.378, 0.012), and negatively related to poverty ratio (-1.9, 95% CI: -3.94, 0.141). Patient activation (PAM-10) was positively related to social support for DSMS (coef = 0.172, 95% CI: (0.105, 0.238)) but only slightly positive for CCM (coef = 0.0526, 95% CI: - 0.0205, 0.126). Lastly, patient activation was positively related to CAHPS G Trust for DSMS (coef = 3.67, 95% CI: 2.18, 5.15) but not related for CCM (coef = -0.70, 95% CI: -2.63, 1.23).

Fig 4. **Patient Activation Measure (PAM-10) primary contrast over time.**. (See Fig 3 for panel descriptions.) All three contrasts overlap 0; thus there is no statistically significant difference between them. DSMS, diabetes self-management support empowerment model; CCM, chronic care model, PAM-10, Patient Activation Measure.

**Table 11.**
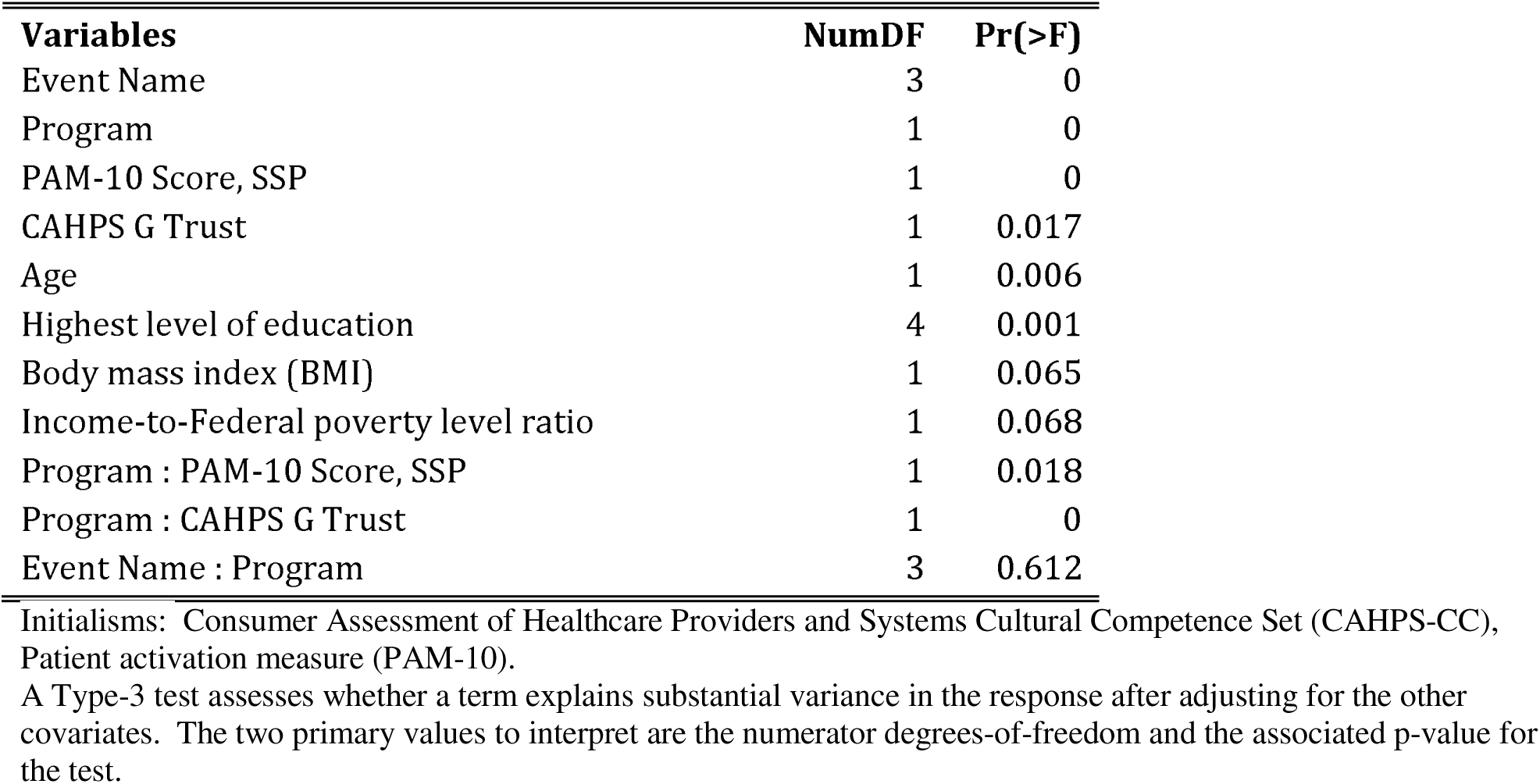
PAM-10 outcome Type III ANOVA Table for reduced longitudinal model.

#### Aim #2: Secondary outcomes

Our second aim was to measure and compare PP success at diabetes self-management as indicated by improvement at 6 months in A1c, depression index score, and body mass index (BMI).

#### Changes in A1c

A1c on the log_2_ scale (log_2_ (A1c), here called “A1c”) had an initial slight decrease for patient participants in both programs (Fig 5A^2^), and the A1c distribution was hyper-exponential showing right skewness even after log transformation. Model fit assumptions were not violated. There was no Time-by-Program interaction (Fig 5C, p-value = 0.616), so both programs changed to similar extents over time; in particular, the Baseline-to-6-month change for CCM compared to the change for DSMS was -0.0165, 95% CI (-0.0786, 0.0456). DSMS was slightly higher than CCM for the study period. Both sites decreased their A1c from baseline to 3 months and sustained their improvement throughout the end of the study (Fig 5B).

Fig 5. **Analysis of A1c measures**. A1c longitudinal data and primary contrast over time between programs. (See Fig 3 for panel descriptions.) DSMS, diabetes self-management support empowerment model; CCM, chronic care model; A1c, meaure of glycoylated hemoglobin proportional to chronic glucose levels.

The multivariate analysis (Table 12, contrasts not shown) revealed that A1c was positively related to depression (coef = 0.0204, 95% CI: (-0.00129, 0.0422)). A1c was positively related to PP DKQ-23 Score at baseline, but much less so at follow-up, and negatively related to SSP DKQ-23 score at baseline, with slightly positive relationships at 3 months and beyond. Males started with higher A1c than females at baseline (2.99 vs 2.89) but both reduced to similar levels sustained from 3 months (2.84), negatively related to age for DSMS (coef = -0.008, 95% CI: (-0.0132, -0.00279)), neutral for CCM (coef = -0.000921, 95% CI: (-0.00745, 0.00561)), higher for females at DSMS than at CCM (diff = 0.14, p-value = 0.0123), but for males about the same between programs. A1c was strongly negatively related to BMI at baseline (coef = -0.0128, 95% CI: (-0.0198, -0.00578)) than at the 3-, 6-, and 12-month follow-ups (coefs = -0.0060, -0.0070, and -0.0059, respectively), and negatively related to BMI at the CCM site (coef = - 0.0147, 95% CI: (-0.0242, -0.00516)) but neutral at the DSMS site (coef = -0.00117, 95% CI: (-0.00988, 0.00753)).

**Table 12.** A1c outcome Type III ANOVA Table for reduced longitudinal model.

| Variables | NumDF | Pr(>F) |
| --- | --- | --- |
| Event Name | 3 | 0.008 |
| Program | 1 | 0.902 |
| DKQ-23 Score, social support participant | 1 | 0.797 |
| Age | 1 | 0.037 |
| Gender | 1 | 0.637 |
| Body mass index (BMI) | 1 | 0.016 |
| DKQ-23 Score | 1 | 0.150 |
| Depression, log2(PHQ-9 + 2) | 1 | 0.065 |
| Event Name : DKQ-23 Score | 3 | 0.061 |
| Event Name : DKQ-23 Score, SS | 3 | 0.014 |
| Event Name : Gender | 3 | 0.021 |
| Event Name : Body mass index (BMI) | 3 | 0.015 |
| Program : Age | 1 | 0.096 |
| Program : Gender | 1 | 0.041 |
| Program : Body mass index (BMI) | 1 | 0.040 |
| Event Name : Program | 3 | 0.229 |

Initialisms: Diabetes knowledge questionnaire (DKQ-23), Depression severity (PHQ-9).
A Type-3 test assesses whether a term explains substantial variance in the response after adjusting for the other covariates. The two primary values to interpret are the numerator degrees-of-freedom and the associated p-value for the test.

#### Changes in Depression

Overall, depression as measured by the PHQ-9 and transformed as log2(PHQ-9 + 2), showed an initial decrease from baseline to 3 months which sustained over time (Fig 6A^3^). Model fit assumptions were not violated. There was a Time-by-Program interaction (p-value = 0.006) because the degree of change over time differed between programs (Fig 6C); in particular, the Baseline-to-6-month change for CCM compared to the change for DSMS was -0.266, 95% CI (-0.464, -0.0669). CCM started lower than DSMS and showed a large initial decrease and continued downward throughout the study while DSMS showed a small initial decrease through

6 months, then started increasing again by 12 months (Fig 6B), nearly rebounding to their baseline levels.

Fig 6. **Depression scores constrasted between treatment groups.** Depression (PHQ-9) longitudinal data and primary contrast over time between programs. (See Fig 3 for panel descriptions.) DSMS, diabetes self-management support empowerment model; CCM, chronic care model; PHQ-9, Depression scores.

The multivariate analysis (Table 13, contrasts not shown) revealed that depression was negatively related to the *Consumer Assessment of Healthcare Providers and Systems Cultural Competence Set* (CAHPS-CC) A and B Communication (coef = -0.175, 95% CI: (-0.29, -0.0605)), negatively related to CAHPS G Trust (coef = -0.117, 95% CI: (-0.23, -0.0047)), and negatively related to Patient PAM-10 Score (coef = -0.0103, 95% CI: (-0.0145, -0.00605)) varying slightly over time. Depression was negatively related to social support PAM-10 score which describes the social support’s perception of the patient participants activation, with the strongest negative relationship at baseline but positive at 6 months, positively related to age at 6 months and negatively related at baseline and 12 months. Depression was negatively related to patient participant PAM-10 Score for DSMS (coef = -0.0068, 95% CI: (-0.0112, -0.00243)), but neutral for CCM (coef = 0.00156, 95% CI: (-0.00319, 0.00631)), and negatively related to income-to-federal poverty level ratio for DSMS (coef = -0.295, 95% CI: (-0.488, -0.102)) but positively related for CCM (coef = 0.196, 95% CI: (-0.0966, 0.489)).

**Table 13.**
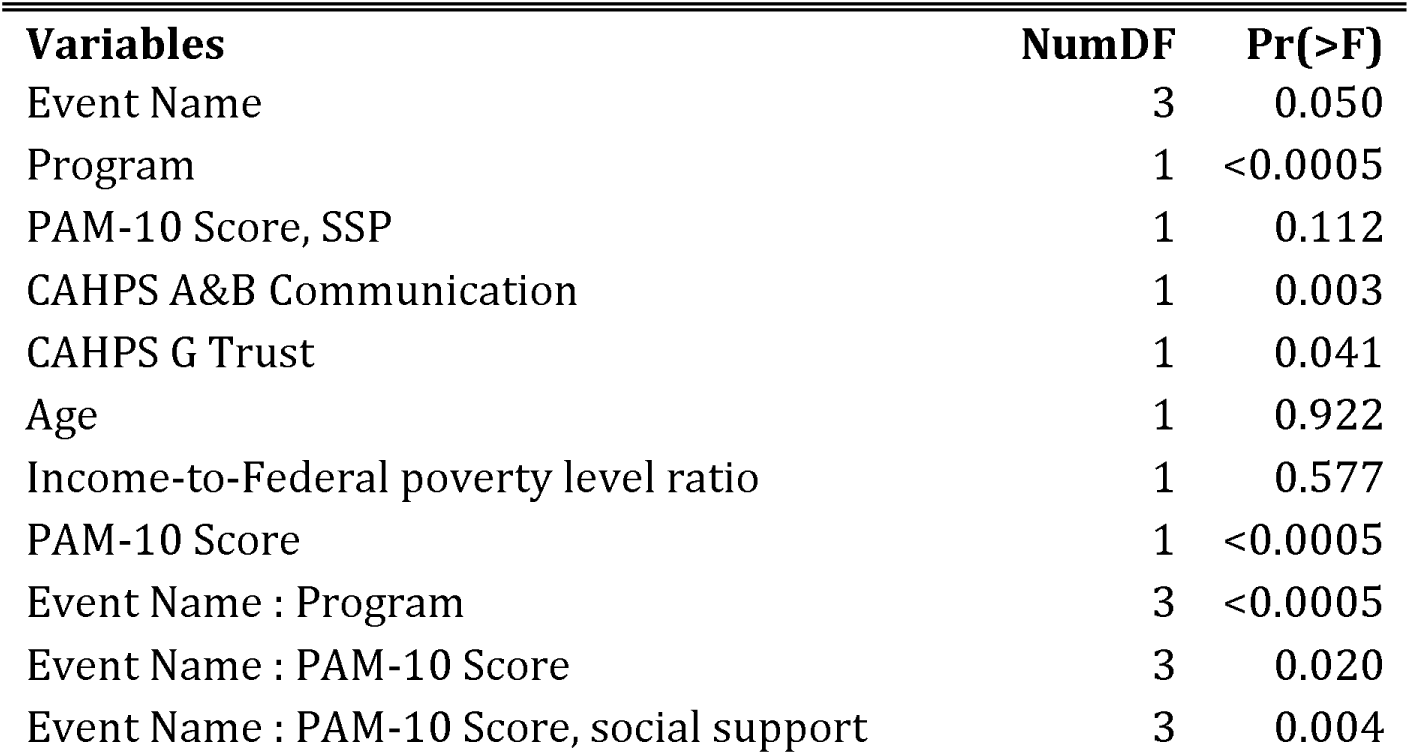
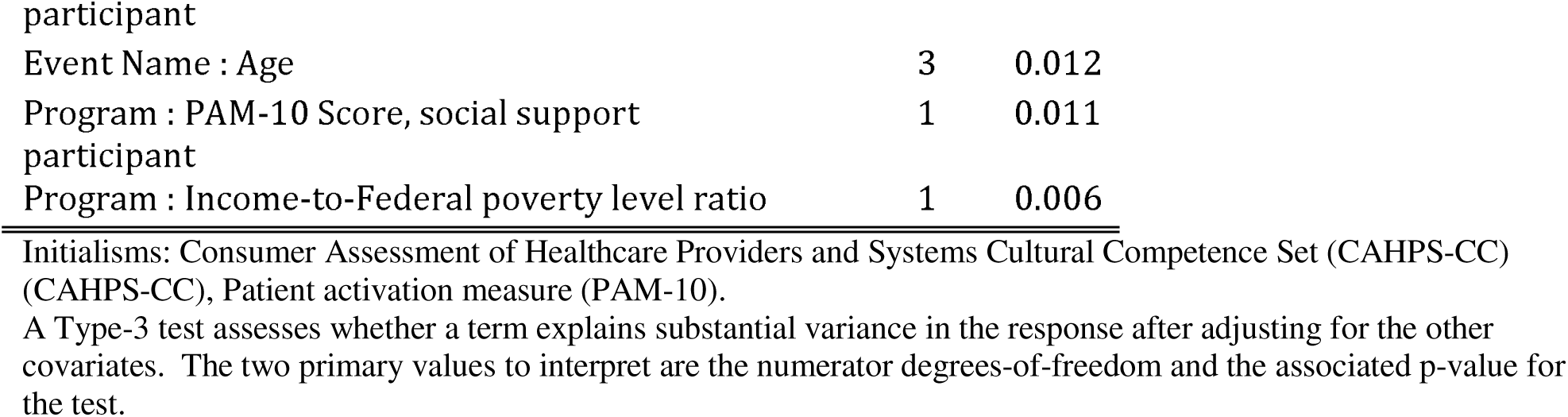
Depression (PHQ-9) outcome Type III ANOVA Table for reduced longitudinal model.

#### Changes in Body Mass Index (BMI)

BMI values were very consistent over time (Fig 7A). Model fit assumptions were not violated. There was no Time-by-Program interaction (Fig 7C, p-value = 0.620), so patient participants in both programs changed to a similar extent over time; in particular, the Baseline-to-6-month change for CCM compared to the change for DSMS was -0.148, 95% CI (-0.204, 0.501). DSMS had slightly higher mean BMI (difference=1.271 BMI) than CCM across the study period Fig 7B).

Fig 7. **Analysis of body mass index (BMI) over time and comparison between cohorts.** BMI outcome longitudinal model data and primary contrast over time between programs. (See Fig 3 for panel descriptions.) DSMS, diabetes self-management support empowerment model; CCM, chronic care model; BMI, body mass index.

A multivariate analysis (Table 14, contrasts not shown) revealed that BMI was negatively related to age (coef = -0.0844, 95% CI: (-0.157, -0.011)) with the effect only slightly varying over time, and positively related to PP DKQ-23 Score (coef = 0.0481, 95% CI: (0.00373, 0.0925)). BMI was positively related to depression for DSMS (coef = 0.17, 95% CI: (-0.0142, 0.357)) but neutral for CCM (coef = -0.0402, 95% CI: (-0.238, 0.157)).

**Table 14.** BMI outcome Type III ANOVA Table for reduced longitudinal model.

| Variables | NumDF | Pr(>F) |
| --- | --- | --- |
| Event Name | 3 | 0.041 |
| Program | 1 | 0.386 |
| Age | 1 | 0.024 |
| DKQ-23 Score | 1 | 0.034 |
| Depression, log2(PHQ-9 + 2) | 1 | 0.343 |
| Program : Depression, log2(PHQ-9 + 2) | 1 | 0.124 |
| Event Name : Age | 3 | 0.026 |
| Event Name : Program | 3 | 0.620 |

Initialisms: Diabetes knowledge questionnaire (DKQ-23), Depression severity (PHQ-9), Body mass index (BMI). A Type-3 test assesses whether a term explains substantial variance in the response after adjusting for the other covariates. The two primary values to interpret are the numerator degrees-of-freedom and the associated p-value for the test.

#### Subpopulation considerations: Results of heterogeneity of treatment effect

Heterogeneity of Treatment Effect was assessed with the same longitudinal model structure with a time-by-program interaction but included only covariates for language, gender, and poverty and their interactions with each time and program.

Associations included DKQ and PAM with language, A1c with gender, and BMI with poverty, but depression had no associations. DKQ-23 for patient participants speaking “English, only” and “Spanish and English” were both about 1.8 points higher than “Spanish, only” (roughly, 15.7 vs 13.9). PAM-10 for patient participants speaking “Spanish, only” (coef = 83.6, 95% CI: (81.4, 85.8)) was about 5 points higher than “English, only” (coef = 78.7, 95% CI: (74.8, 82.6)), with “Spanish and English” (coef = 81.4, 95% CI: (77.8, 85)) being in the middle and not statistically different from either “only” category.

A1c at baseline for males (coef = 2.99, 95% CI: (2.89, 3.08)) was higher than females (2.9, 95% CI: (2.84, 2.96)), but both genders had similar A1c values from 3 months through 12 months (roughly, coef = 2.83, 95% CI: (2.73, 2.93)). A1c for females at DSMS (coef = 2.91, 95% CI: (2.84, 2.98)) was higher than at CCM (coef = 2.78, 95% CI: (2.7, 2.86)), but males at both programs had similar A1c values (roughly, coef = 2.88, 95% CI: (2.74, 3.00)).

PHQ-9 was not related to poverty, gender, or language, only to program and time. BMI was higher for patient participants above the FPL at DSMS (coef = 34.3, 95% CI: (32.6, 35.9)) than at CCM (coef = CCM = 30.2, 95% CI: (27.1, 33.4)), but about the same between programs for patient participants below the FPL (both roughly coef = 32.8, 95% CI: (31.5, 34.2)).

Outcome changes by baseline PAM, A1c, PHQ-9, and BMI categories are grouped by their baseline categories and plotted over time to show differential effects in Fig 8^4^. These analyses should be interpreted cautiously because of the small sample sizes.

PAM overall trends showed a slight increase for both programs, but there are larger increases in the few patient participants who were lowest at baseline with very significant and greater change at CCM. There was no change for those patient participants who were already activated in either program, which represented the vast majority.

A1c remained consistently low in both programs for prediabetes and diabetes patient participants who started the program with lower A1c levels (<9). Both programs had large effects on higher A1c levels (≥9), with the CCM program showing the strongest effect. Neither program resulted in A1c reaching normal glycemic levels of <5.6, but at CCM high A1cs dropped below 9, which is clinically meaningful. Both programs kept those with prediabetes from elevating into the diabetes range and kept lower diabetes range scores from elevating higher. This is also clinically meaningful. While CCM patient participants, even those entering at the higher levels, dropped below 9, the higher A1c group in the DSMS program did not achieve that degree of diabetes control. The largest effect was seen at the 3-month time point in both programs.

PHQ-9 depression scores showed the most difference between baseline and subsequent time points in CCM, with some smaller improvements in DSMS. Again, the biggest change was in the first three months with some continuing improvement at 6 months. The least depressed group did not improve further.

BMI was static in both programs, remaining approximately constant at baseline and at 12 months, (32.6 and 33.6 respectively). PAM-10 trajectories are similar between programs (Fig 9). Patient participants in the highest-A1c baseline category at CCM had greater decreases in A1c than at DSMS, with more than half of them lowering below 9 (Fig 10^5^). PHQ-9 shows that depression is higher with Baseline A1c, at Program DSMS depression is higher than at CCM overall, decreases at CCM are greater at all A1c baseline categories with median PHQ-9 values ending below 5 for all categories, while DSMS ends below 5 in only the lowest A1c category and all categories showing increases in depression by 12 months (Fig 11^6^). BMI trajectories are similar between programs with no interpretable pattern (Fig 12).

Fig 8. **Comparison across four outcome measures (PAM-10, A1c, PHQ-9, and BMI).** The mean change over time for each group is defined by their baseline group category. PAM-10 levels: 0.0 - 45.1, Level 1 Disengaged and overwhelmed; 47.4 - 52.9, Level 2 Becoming aware, but still struggling; 56.0 - 72.1, Level 3 Taking action; and 75.5 - 100, Level 4 Maintaining behaviours and pushing further. A1c levels: 0 - 5.7, Neither; 5.7 - 6.4, Pre-diabetes; and 6.4 - 14+, Diabetes. PHQ-9 levels: 0 - 4, None-minimal; 5 - 9, Mild; 10 - 14, Moderate; 15 - 19, Moderately Severe; and 20 - 27, Severe. BMI levels: 0 - 18.5, Underweight; 18.5 - 25, Normal or Healthy Weight; 25 - 30, Overweight; and 30+, Obese. DKQ Score has no defined levels so is not shown. DSMS, diabetes self-management support empowerment model; CCM, chronic care model; PAM-10, Patient activation measure; PHQ-9, Depression severity (PHQ-9); BMI, body mass index.

Fig 9. **PAM-10 outcome by A1c Baseline category**. Small improvement in most A1c catagories occurs in both groups in the first 3 months and in the lowest category over the year. Numbers of subjects in each A1c category for each cohort appears above the trajectory. (See Fig 8 for categories.). DSMS, diabetes self-management support empowerment model; CCM, chronic care model; PAM-10, Patient activation measure.

Fig 10. **A1c outcome by A1c Baseline category.** Patient participants in the highest-A1c baseline category at CCM had greater decreases in A1c than at DSMS, with more than half of them lowering below 9. (See Fig 8 for categories.) DSMS, diabetes self-management support empowerment model; CCM, chronic care model.

Fig 11. **PHQ-9 outcome by A1c Baseline category.** Depression is higher with Baseline A1c, decreases at CCM are greater at all A1c baseline categories. (See Fig 8 for categories.) DSMS, diabetes self-management support empowerment model; CCM, chronic care model; Depression severity (PHQ-9).

Fig 12. **BMI outcome by A1c Baseline category.**BMI trajectories are similar between programs. (See Fig 8 for categories.) DSMS, diabetes self-management support empowerment model; CCM, chronic care model; BMI, body mass index.

### Programmatic Analyses

#### Aim #3: Culture and context of programs

Our third aim was to characterize the ways that two distinct culturally competent diabetes self-management programs (DSMS and CCM) interface with patient participant culture and socioeconomic context.

#### Program assessments

The section on “comparators” provides a description of each program. Both emphasize the importance of understanding patient needs, including from a cultural and contextual perspective. One staff member at DSMS said that they do not just talk about what to eat or not eat—they cover depression, stress management, and difficult topics like erectile dysfunction, kidney failure, and amputation. They do a meditation and deep breathing exercise to help people learn alternative methods for stress reduction, they encourage the patients to bring a social support with them to the classes (without charge), and they were in the process of developing a relationship with the local food bank in order to better address patient needs in relations to diet and food insecurity. A strength of the DSMS program from the perspective of program staff is their emphasis on identifying individual needs and tailoring the experience for each patient with a heavy focus on helping people learn how to set goals. The group dynamic of the classes was identified as a strength. Challenges identified primarily related to lack of time, insufficient staff, space, funding, and resources. The DSMS program made some alterations to the way they delivered the program during the middle of the project. These changes were required by guidance from the ADA to maintain program accreditation.

The strengths of the CCM program from the perspective of staff are the relationship of trust that they enjoy in the community, the fact that the staff are both “in” and “of” the community, and that the services provided are holistic and wrap-around. The challenges identified were lack of financial resources and a need for more staff and space. At CCM, because the program is community-led and community-run, the program focuses holistically on the patient and their family, and the CCM facility has a feel more like somebody’s home than that of an office or a clinic. A broad range of services is available, in addition to the ongoing diabetes classes, including primary care, dental care, psychiatric care, a food coop, children’s educational activities and classes, and exercise classes. CCM is operated by a faith-based nonprofit, so the spiritual dimension of people’s lives is incorporated if the patient would like to “sit and pray” alone or with others. Stress management is covered in the diabetes classes, and mental and behavioral health services are available. Through a number of CHW-run programs on site, CCM screens patients for social needs and provides navigation, including related to domestic violence, legal services, education, parenting, substance misuse, housing insecurity, food insecurity, and access to safety net programs like SNAP, WIC, prescription discount programs, utilities support programs, and Medicaid. CCM does not accept insurance, and the fee for services is nominal or waived, depending on patient circumstance and capacity to pay.

A primary difference between the two sites is in relation to language—a core element of program cultural competence. At DSMS, the default program language spoken is English, but approximately 25% of patients are Spanish speakers, with a smaller number monolingual Spanish speakers. All of the handouts DSMS uses are available in Spanish, including recipes, but only one staff person speaks any Spanish. A primary concern among DSMS staff was the inability of the program to hire a Spanish-speaking diabetes educator to lead the classes because of budgetary constraints. Monolingual Spanish-speaking diabetes patients are given a truncated delivery of the class using an interpreter.

The default language at CCM is Spanish but there is significant capacity for bilingual communication. The vast majority of CCM patients are Spanish-speaking—either monolingual or bilingual. All of the staff at CCM are Spanish-speaking, and most are bilingual in Spanish and English. A few staff are monolingual Spanish speakers. Volunteer medical providers who see patients at CCM (primarily MDs from UNM) and volunteer educators (UNM pharmacy students, medical students, and family medicine residents) are a mix of bilingual and non-Spanish-speaking. However, a translator is used if needed.

#### Cultural competence surveys

Consumer Assessment of Healthcare Providers and Systems Cultural Competence Set (CAHPS-CC) composite scores for five domains had a similar pattern between programs. Overall quality of care, communication, equitable treatment, and trust were rated very high by participants in both programs. Overall quality of care (A), Patient Participant-Provider Communication (B), and Trust (G) were scored higher by patient participants at CCM than at DSMS. Interpreter Services (H) was scored slightly lower among patient participants who required interpreter services, but more negative for patient participants at DSMS than at CCM (Table 15).

**Table 15.** Consumer Assessment of Healthcare Providers and Systems Cultural Competence Set (CAHPS-CC) Cultural Competence domain scores.

| CAHPS Cultural Competence | (N=) | All Patient Participants<br>(N=226)<br>Median [Q1; Q3] | Program |  | P<sup>a</sup> |
| --- | --- | --- | --- | --- | --- |
|  |  |  | CCM<br>(N=106)<br>Median [Q1; Q3] | DSMS<br>(N=120)<br>Median [Q1; Q3] |  |
| PP and SSP, quality of care | 216 | 0.86 [0.74;0.93] | 0.87 [0.78;0.94] | 0.83 [0.71;0.92] | 0.030 |
| A&B. PP-Provider Communication | 217 | 0.96 [0.90;1.00] | 0.97 [0.93;1.00] | 0.95 [0.85;1.00] | 0.007 |
| F. Equitable treatment | 217 | 1.00 [1.00;1.00] | 1.00 [1.00;1.00] | 1.00 [1.00;1.00] | 0.064 |
| G. Trust | 217 | 0.90 [0.74;0.97] | 0.92 [0.79;0.97] | 0.88 [0.67;0.96] | 0.050 |
| H. Interpreter Services | 82 | -0.11 [-0.22;0.00] | -0.06 [-0.20;0.00] | -0.13 [-0.23;-0.11] | 0.008 |

<sup>a</sup>P-values reported from the Kruskal-Wallis test for continuous and from the chi-square test with continuity correction for categorical data.
Initialisms: Diabetes Self-management support empowerment model (DSMS), Chronic care model (CCM), Consumer Assessment of Healthcare Providers and Systems Cultural Competence Set (CAHPS-CC).

## Discussion

Our hypothesis was that diabetes self-management programs are most successful if their design is culturally and contextually “situated” [40,43,44] by positively leveraging the cultural values and accommodating the socio-economic circumstances of a patient population in a way that creates synergy with patients’ everyday lives [20–26,29,30,81]. To test this hypothesis, we compared two program models--one based in an academic medical center using an approach based on group educational sessions taught by a trained diabetes educator and the other, a community-run program based on a wrap-around services model. Although they are both described as “culturally competent,” they differ strongly in the extent to which they embody characteristics of cultural and contextual situatedness.

### Primary outcomes: Diabetes knowledge and patient activation

For diabetes knowledge and patient activation (capacity for change), we found no statistical differences between programs.

Diabetes knowledge increased slightly in patient participants in both programs. DSMS patients started higher but the change at both sites was parallel. At DSMS, better “patient participant-provider communication” as measured by the Consumer Assessment of Healthcare Providers and Systems Cultural Competence Set (CAHPS-CC), was associated with higher diabetes knowledge scores. At CCM, there was a counter-intuitive association between less effective patient participant-provider communication and higher diabetes knowledge scores, which on face value was challenging to interpret.

Most patient participants at both sites reported high capacity for diabetes self-management and scored highly on our patient activation measure, which made it difficult to show any differences (ceiling effect). Both programs were generally similarly effective at increasing a patient participant’s activation from baseline to three months and tended to stay at the higher level for the rest of the study period.

However, there were differences between the sites, with CCM patient participants generally being more activated than DSMS patient participants. We believe that the higher-level activation at CCM is related, in part, to the need for CCM participants to rely on themselves rather than medical treatment for managing their diabetes or their health in general because, as indicated above, few CCM patients have health insurance. Moreover, in the interviews and focus groups, we gathered data suggesting that few CCM patient participants qualify for safety net benefit programs that provide supports for unemployment, disability, food insecurity, or rental assistance. Most patient participants at DSMS were using these safety net programs, including many on Social Security Disability Insurance (SSI). Therefore, as indicated in the discussion of the qualitative results, we found that CCM patient participants are generally much more concerned than DSMS patient participants about the potential of diabetes to negatively impact their ability to work and to provide support for a family—and more patient participants at CCM were married or living with a partner than at DSMS.

In addition, patient participants in the CCM program who scored very low on the PAM at baseline made more and sustained change, ending up as high or higher than those with higher scores at both time points (please interpret this subgroup observation with caution). Those with low activation at DSMS were not able to become activated to the level of the vast majority of other participants in the study.

### Secondary outcomes: changes in A1c, BMI, and depression

For our secondary outcomes of changes in A1c, and BMI, we did not find statistically significant differences between programs. In relation to A1c, we found that blood sugar levels at both sites decreased and that the difference in the decrease between the sites was not statistically significant. At both sites, after initial decrease at three months, the change was sustained for the rest of the study—although the change was not statistically significant at either site. The decrease seems slightly better sustained at DSMS, but the actual A1c values began lower and stayed lower at CCM.

Baseline values at both sites were similar in relation to the number of patient participant A1cs indicating prediabetes (6 vs 8), low (39 vs 41), medium (36 vs 31), and very high (13 vs 15) A1c values, but DSMS had notably more patient participants in the high range (26 vs 11).

Although neither program lowered participant A1c into a non-diabetes range, patient participants with prediabetes in both programs had A1c scores that remained below 6.4 and lower diabetes range scores (6.4-9.0) did not elevate higher. While stable A1c levels could be due to a variety of factors, and prediabetes does not inevitably get worse, especially over a short period of observation, this is a desired clinical outcome with long-term health implications. The literature demonstrates [82–86] that every 1% decrease in A1c is associated with improved health outcomes and reduced risk related to developing complications such as retinopathy/blindness, kidney disease, neuropathy, diabetes-related hospitalization, cardiovascular disease (CVD), and diabetes-related mortality. Skyler and colleagues [82] found that for each 1% decrease in A1c, there was a statistically significant 18% reduction in CVD events—the primary risk of death for people with diabetes. Similarly, Huang and colleagues [87] found that risk of complications and death became significantly higher above 9.0. The risk threshold for diabetic ketoacidosis (“diabetic coma”) [88] is also an A1c of 9. Therefore, of greater interest, although the difference in the change in A1c over time was not found to be statistically significant between programs, nor was the change dramatic, there was a key clinically meaningful difference, especially true for patient participants with higher baseline A1c values. At CCM, average “very high” A1cs dropped below 10 and the median was below 9. While on average CCM PPs, even those entering at the higher levels, dropped below 10—and the median was below 9, the higher A1c group in the DSMS program did not achieve that degree of diabetes control. The results for patient participants who had high A1c at baseline and dropped on average below 10 with the median below 9 have positive health implications.

Patient participants at CCM, perhaps in part because they tended to start with lower A1c values overall (including fewer with A1c over 10), were more activated [89], and were less depressed (discussed below) at baseline, were able to bring their A1c down to approximately 9.0, the risk threshold for diabetic coma [88]. At DSMS, in part because they started with higher A1cs, were less activated, and more depressed (discussed below), patient participants tended not to lower their A1c near 9.0. Because of the clinically important impact of each 1% increase/decrease in A1c on risk for diabetes-related health complications and the increased risk of complications over time, projected long-term clinical outcomes for the two sites would appear to be meaningfully different. Because more change and a sustained lower level was obtained by CCM patient participants, CCM came much closer to achieving the goal of managing their diabetes than patient participants at DSMS.

There was no difference between the sites in relation to improvement in BMI. Neither site produced reductions.

An important area where we found a statistically and clinically significant difference between the two sites and also revealed important dynamics related to social support was in relation to depression symptoms [72]. CCM started with baseline depression scores that were generally lower than those at DSMS, though at both sites a majority of participants had either “none-minimal” or “mild” severity scores (CCM 84.9%, DSMS 71.7%). Both sites reduced their level of depression symptoms after baseline, but despite starting with less depression (which one could hypothesize should make it more difficult to change), depression scores at CCM improved more than DSMS, and while CCM improvement continued over the period of the study, at DSMS, the improvement was not sustained. Again, the biggest change was in the first three months with some continuing improvement at 6 months, except for the most highly scoring patient participant in the CCM program, where improvement appeared only at 6 months. The least depressed group did not improve further.

As anticipated from research on diabetes and depression, we found that depression symptoms were related to A1c. Patient participants with higher depression scores tended to have higher A1cs. We also found that the diabetes knowledge of the social support was important— higher diabetes knowledge of the social support was associated with lower depression symptoms and better A1c values. The design of the CCM program that involves ongoing peer support together with the demonstrated high level of trust and cultural competence at CCM can be hypothesized to provide a more “supportive” environment involving more in-person contact and the creation of a larger social network. We hypothesize that more trust in the site as indicated in scores for the Consumer Assessment of Healthcare Providers and Systems Cultural Competence Set (CAHPS-CC) (Table S 1) may lead participants to be more interested in participating and develop a feeling of commitment to others related to “showing up” to ongoing group and individual meetings. These meetings become a type of peer support, with patient participants who attend sharing recipes and strategies for diabetes management. Our interpretation is that this sharing and empathy likely contribute to the lower and more sustained levels of depression symptoms. Participants at both sites indicated the importance of these factors in qualitative data, and our broader research with Latina and Mexican immigrant women and CHWs in Albuquerque strongly supports this finding of the deep importance of social support in the lives of Latinxs [90–95]. In fact, this is why our patient advisors were adamant that we find a way to include social support and social context in the design of our study, and why we enrolled participants as patient participant-social support participant dyads rather than merely recruiting patient participants to participate as individuals.

Furthermore, although patient participants at both sites rated their program high in cultural competence and expressed that they really like the program and the staff, when posed with a question about what was missing from the program design, DSMS patient participants tended to indicate that the DSMS model of six discrete group sessions was not really sufficient and was less attractive than a program like the CCM that provides ongoing classes and support. DSMS patient participants and their social support participants reported that patient participants often feel socially isolated and alone in relation to the issue of diabetes—and CCM patient participants discussed the importance of having the social support of the program in an ongoing way. Greater social support and interaction and less social isolation at CCM might help to explain the lower levels of depression symptoms overall, as well as the greater and more sustained decrease in depression after joining the program.

Data modeling and visualization revealed that changes were not uniform, thus the exploratory, post-hoc subgroup analysis helped provide additional understanding of who had bigger changes than others. A subgroup analysis by A1c baseline category showed that depression symptoms at DSMS is higher than at CCM overall, decreases at CCM were greater at all A1c baseline categories with median values ending below 5 for all categories, while DSMS ends below 5 in only the lowest A1c category and all categories showing increases in depression by 12 months.

### Cultural competence of programs

Overall Consumer Assessment of Healthcare Providers and Systems Cultural Competence Set (CAHPS-CC) composite scores were high at both sites. Nearly all patient participants rated their program positively, and the range was not wide. CCM was somewhat more positive. The CAHPS-CC sub-scales reflect domains that have been identified as important components for culturally competent programming. Two key CAHPS-CC sub-scales did demonstrate statistically significant differences between the sites: “Patient participant-provider communication” and “Trust”. CCM scored very high for both domains. We hypothesize that these two characteristics are associated with cultural competence. Our take-away from these results is that CCM outperformed DSMS on the CAHPS-CC. While both sites do an excellent job addressing issues of cultural competence through program design and services, CCM is the more culturally competent of the two sites—a finding that aligns with both our theoretical model and with the discussion and interpretation of the outcomes below. The CCM is designed to address the specific needs of Latinx patients from low-income households [29,30] by creating comprehensive, integrated, wrap-around services focused on culturally competent care].

### Outcomes and Comparative Effectiveness of the Two Programs

Findings from this study demonstrate comparative effectiveness of two culturally and contextually situated models of diabetes self-management and education. The two models were similar in many ways: both were deemed culturally competent by PPs and SSPs, and we did not find statistically different results between the two sites with respect to the primary outcome measures and only one of the three secondary outcomes showed differential results. Moreover, the design of this study was observational, and given major differences in site context and patients enrolled, residual confounding is likely. However, given the higher cultural competence rating (0.87 vs 0.83), statistically significant improvement in depression (larger decrease by - 0.266) [72], and the importance of social support to the patients (84.9% vs 71.4% strongly agree), the results suggest that culturally and contextually situating a diabetes intervention may deliver benefit for patients, especially for some subgroups of patients.

While the results appear to align with findings in the broader literature that cultural competence plays a role as beneficial for health promotion programming [27,81], especially for some populations, we recognize that the CCM model requires a substantial commitment to creating wrap-around services that put the patient and the patient’s needs at the center of care and include taking values, customs, beliefs, and language from Latinx culture into account. This is not a model currently in wide use, and it could be challenging for some sites to implement. While it was not within the scope of this study to speak further to the generalizability of the model, further study may show the more nuanced ways that each of these models operates to produce cultural competence for sub-populations.

### Understanding the results

Comparison of the two programs and patient participant outcomes at both sites demonstrated less overall difference than hypothesized; however, differences between the two sites in patient participant perceptions of program cultural domains of trust and patient participant-provider communication, depression scores, associations with poverty, and the clinical significance of comparative A1c scores align with our hypothesis of the importance of cultural and contextual situatedness in creating program cultural competence. A culturally and contextually situated program positively leverages the cultural values and accommodates the socio-economic circumstances of a patient population in a way that creates synergy with patients’ everyday lives [20–26,29,30,81].

Strikingly, neither program achieved the goal of reaching an A1c of 7.0, neither program showed change in BMI, and only the CCM program showed better depression scores, although not to a non-depressed state, (PHQ-9 <4). At CCM, depression scores of subjects with both moderately severe or moderate depression (10-14) fell to mild depression (4-9), a meaningful achievement. At DSMS, this improvement was seen in the moderate depression group at 3 and 6 months, but not at the 12-month follow-up. Thus, while there were some advantages to the CCM program, neither program achieved the ultimate goal and made only moderate progress towards the clinical end points of normal glycemia, BMI within normal limits, and no depression. These findings suggest that more work needs to be done to mitigate long term life-threatening consequences of diabetes.

### Limitations and generalizability

We compared two real world programs to avoid the problem of overly controlled or delimited environment that does not replicate real world conditions that can happen with randomized control trials. We recognize that this introduces differences in the populations as well as differences in the interventions and the responses to them that cannot be controlled for in our analysis of the outcomes and that the associations we observed are not necessarily causal.

This study was also limited by variations at intake within each study group. While CCM PPs were patients at a community clinic that does not accept insurance and has nominal fees, those at DSMS were referred by physicians to a program that requires either insurance or self-pay. Differing demographics at intake included income/poverty level, size of households, education level, A1c scores, depression scores, and BMI. Differences in the populations as well as clinical setting could not be adjusted for completely in our multi-variate analyses and may bias the association between self-management programs and the outcomes. And furthermore, data on patient adherence to the DSMS and CCM programs were not available.

This study used a non-randomized quasi-experimental design study. The two comparator sites are distinct in their diabetes management program models, thus allowing (after controlling for other factors) for direct comparison of the effects of the program on the primary and secondary outcomes. This choice of comparators will reduce the potential for bias for the following reasons: (*A)* the two comparators serve relatively similar populations in terms of socio-demographic attributes; *(B)* diabetes self-management program models in use at the two comparator sites have program attributes that are sufficiently distinct to allow contrast and comparison; and, (*C)* each of the comparator sites is implementing a program in a “real life” setting, thus providing the opportunity for a pragmatic assessment of the comparative effectiveness of the program models under externally valid and generalizable conditions.

Patient stakeholder data collectors (PSDCs) were recruited from the patient population. This project could not have been accomplished without this design feature, and the science was significantly enhanced, as evidenced by successful recruitment of a hard-to-reach population and notably low attrition. This finding contributes to the current literature on patient-engaged and community-engaged research and is supported by our own publications.

## Conclusions

Our results suggest that a holistic, culturally and contextually situated approach to diabetes prevention and self-management can be beneficial to patients and to their families, caregivers and social supports. The data suggest that integrated, ongoing support is potentially helpful for patients with diabetes. We learned the important role that social support plays in the lives of diabetes patients and care givers and that more attention needs to be paid to integrating these dimensions of a patient’s reality into care.

This study compared diabetes self-management over a 12-month period for 226 Latinx diabetes patients from two different diabetes self-management programs. Although they are both described as “culturally competent,” they differ strongly in the extent to which cultural characteristics are embedded in their structure and function. The primary outcome, improved capacity for diabetes self-management measured through improvements in diabetes knowledge and diabetes-related patient activation, was equivalent for both programs. Both programs resulted in improved A1c levels, although no change in BMI. The only statistically significant difference between programs was the effect on one of the secondary outcomes of depression scores, which were more improved in the CCM than the DSMS program [72], and were more significant in patients with severe diabetes (A1c > 10). In this A1c catagory, improvement in A1c levels followed decreasing depression scores, indicating a subtle but powerful impact of that treatment model. Decreases of A1c in this range are associated with fewer secondary effects of the hyperglycemia clinically.

Using exploratory, post-hoc subgroup analyses, we found differences between small subgroups of patients based on lower baseline PAM and higher baseline A1c and depression scores, and based on scores on the Consumer Assessment of Healthcare Providers and Systems Cultural Competence Set (CAHPS-CC) that were supported by qualitative data and clinically meaningful differences in A1c outcomes, all of which pointed toward CCM as the more culturally and contextually situated model. In future a study focusing on patients with high A1c levels in a large enough cohort for more statistical power would be useful and may reveal treatment and prevention modalities for the life-threatening sequelae of diabetes. As such, this study demonstrates that culturally and contextually situated approaches can successfully deliver effective benefits for diabetes self-care, and that even more graded interventions may be required to enhance self-management and eliminate the long-term lethal complications of diabetes in this population.

## Data Availability

All UNM-OCH Diabetes Project quantitative data files are available from the University of New Mexico’s Digital Repository https://digitalrepository.unm.edu/hsc_data_othr/1/ [DOI URL]. Qualitative data contain identifiable information and will not be shared.

## Supporting Information

### Human subjects approval

This study was approved by the Human Research Protections Office at the University of New Mexico (#16-303). All participants provided signed informed consent.

### ClinicalTrials.gov registration

This study was registered with ClinicalTrials.gov #NCT03004664.

## Supplement

### Additional questions for Patients

**Table S 1.**
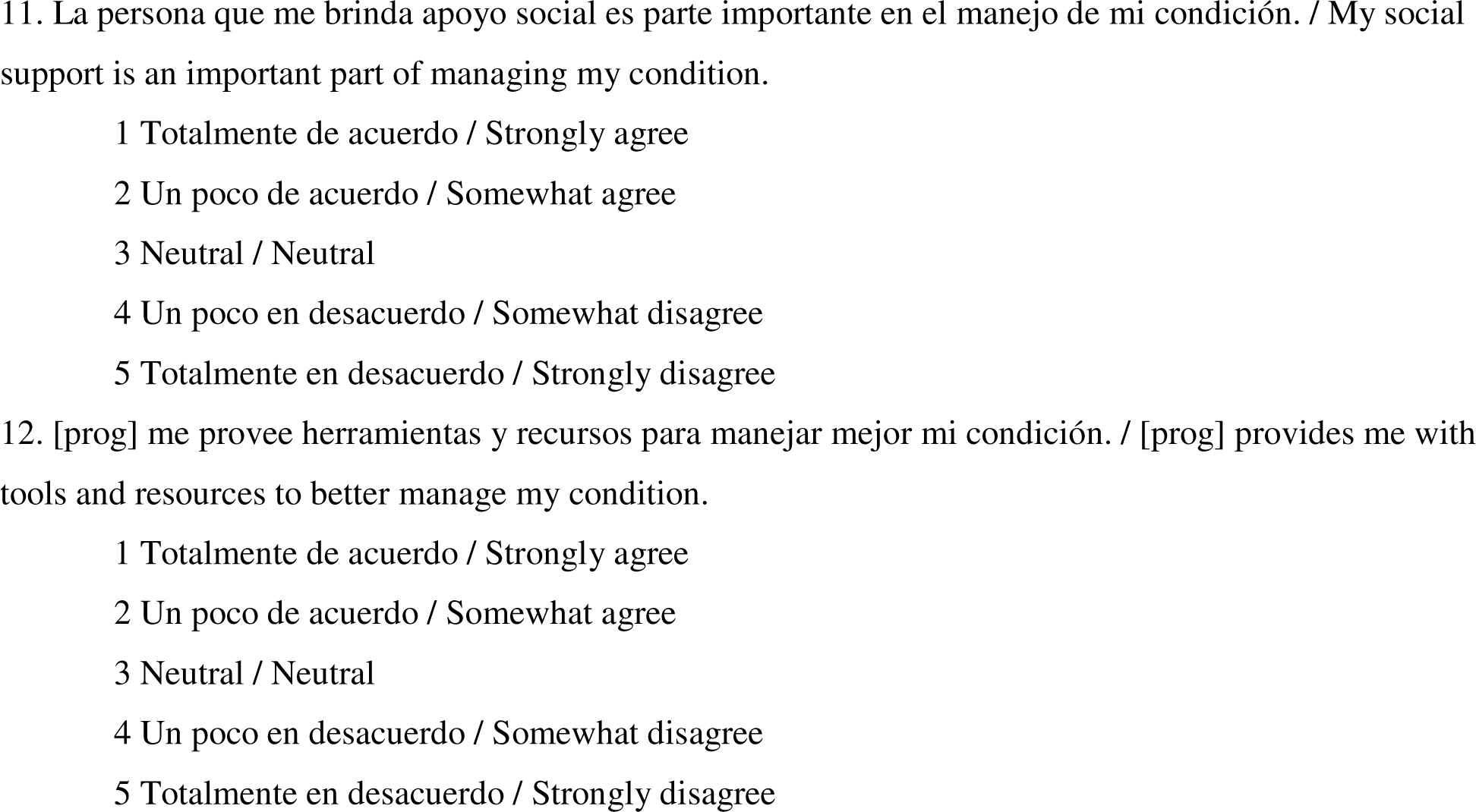
Two added questions to the but were not included in the total scoring.

### Diabetes Knowledge Questionnaire

**Table S 2.**
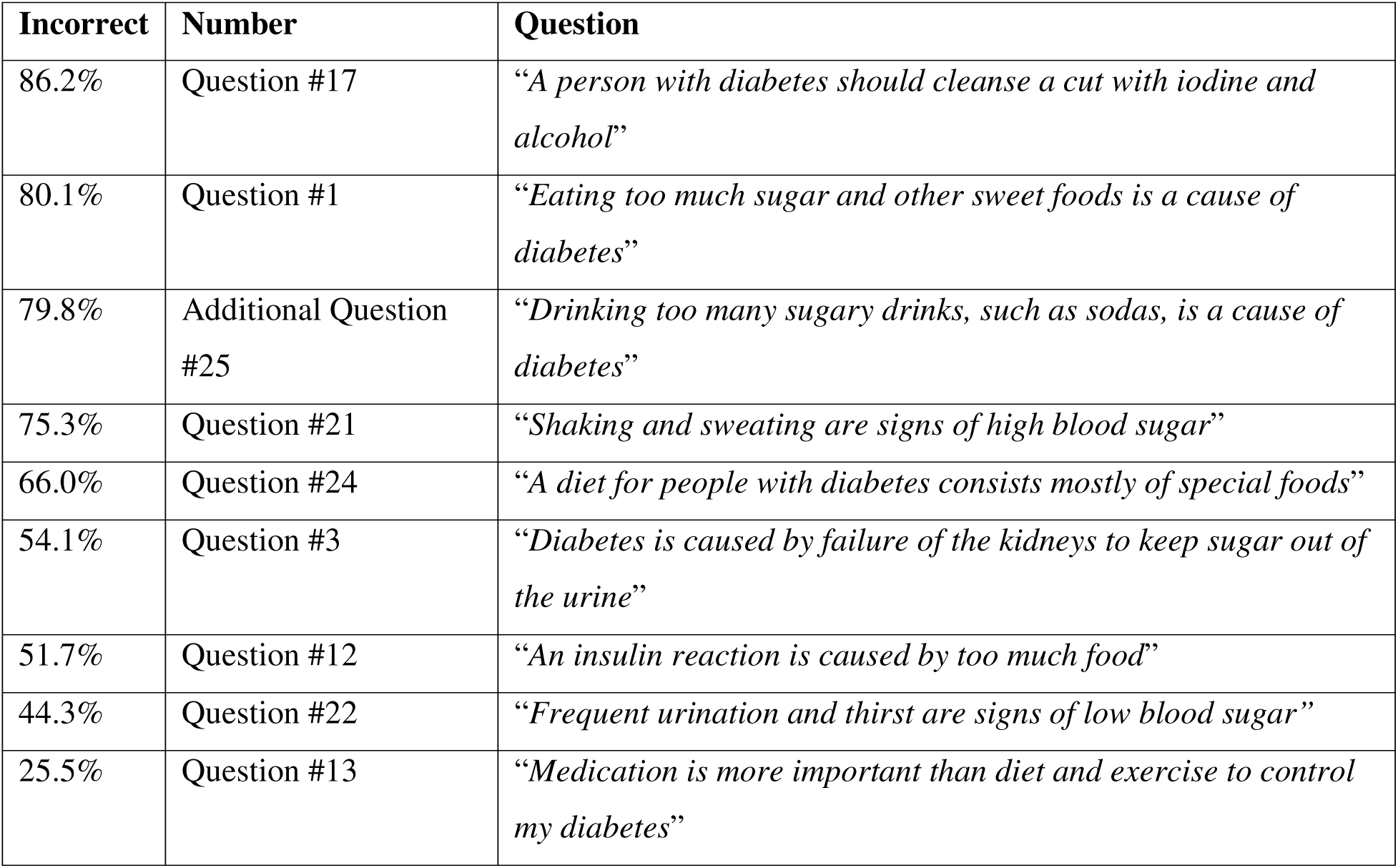
Patients incorrectly answered “yes” to these DKQ questions most often. Interestingly, 100% of all patients correctly answered the question whether people with diabetes should take extra care when cutting their toenails.

| **Incorrect** | **Number** | **Question** |
| --- | --- | --- |
| 86.2% | Question #17 | *“A person with diabetes should cleanse a cut with iodine and alcohol”* |
| 80.1% | Question #1 | *“Eating too much sugar and other sweet foods is a cause of diabetes”* |
| 79.8% | Additional Question #25 | *“Drinking too many sugary drinks, such as sodas, is a cause of diabetes”* |
| 75.3% | Question #21 | *“Shaking and sweating are signs of high blood sugar”* |
| 66.0% | Question #24 | *“A diet for people with diabetes consists mostly of special foods”* |
| 54.1% | Question #3 | *“Diabetes is caused by failure of the kidneys to keep sugar out of the urine”* |
| 51.7% | Question #12 | *“An insulin reaction is caused by too much food”* |
| 44.3% | Question #22 | *“Frequent urination and thirst are signs of low blood sugar”* |
| 25.5% | Question #13 | *“Medication is more important than diet and exercise to control my diabetes”* |

### Patient Activation Measure

**Table S 3.** Patients answered “disagree” or “neutral” to these PAM questions most often (questions are paraphrased so as to conform to the PAM license agreement).

| **Disagree**<br>**or Neutral** | **Number** | **Question** |
| --- | --- | --- |
| 20.5% | Question #4 | Confident about choosing medical attention or self-care for a health issue. |
| 26.9% | Question #7 | Can stick to positive diet and exercise goals. |
| 20.4% | Question #8 | Knowledgeable in preventing health issues. |
| 16.8% | Question #9 | Confident in my ability to find solutions to new health problems. |
| 19.9% | Question #10 | Even during stress, can stick to positive diet and exercise goals. |

## Footnotes

1 This figure was previously published in [72].

2 This figure was previously published in [72].

3 This figure was previously published in [72].

4 This figure was previously published in [72].

5 This figure was previously published in [72].

6 This figure was previously published in [72].

## References

1. Page-Reeves J, Regino L, Murray-Krezan C, Bleecker M, Erhardt E, Burge M, et al. A comparative effectiveness study of two culturally competent models of diabetes self-management programming for Latinos from low-income households. BMC Endocr Disord. 2017 Dec;17(1):1–8.

2. Institute of Medicine (US). Initial National Priorities for Comparative Effectiveness Research [Internet]. Washington, DC: National Academies Press; 2009. Available from: http://www.nap.edu/catalog/12648/initial-national-priorities-for-comparative-effectiveness-research

3. Centers for Disease Control and Prevention. National Diabetes Statistics Report [Internet]. Atlanta, GA: US Department of Health and Human Services; 2022 [cited 2022 Jul 15]. Available from: cdc.gov/diabetes/data/statistics-report/index.html

4. Beckles G, Zhu J, Moonesinghe R. Diabetes - United States, 2004 and 2008. MMWR Morb Mortal Wkly Rep. 2011 Jan 14;60(Suppl):90–3.

5. Office of Minority Health. Diabetes and Hispanic American [Internet]. U.S. Department of Health and Human Services; [cited 2022 Aug 6]. Available from: https://minorityhealth.hhs.gov/omh/browse.aspx?lvl=4&lvlid=63

6. Brown A, Patten E. Hispanics of Mexican Origin in the United States, 2011 [Internet]. Pew Research Center; 2013 Jun [cited 2015 Nov 10]. Available from: https://www.pewresearch.org/hispanic/2013/06/19/hispanics-of-mexican-origin-in-the-united-states-2011/

7. New Mexico Department of Health. Mortality Data, Years 1999-2013 - Leading Causes of Death Counts [Internet]. New Mexico’s Indicator-Based Information System. [cited 2016 Feb 1]. Available from: https://ibis.health.state.nm.us/query/result/mort/MortCnty/LCDCount.html

8. New Mexico Department of Health. New Mexico Prediabetes and Diabetes Facts. 2015 May.

9. Harvard T.H. Chan School of Public Health. Press release: New poll finds diabetes top health concern for Latino families [Internet]. News. 2014 [cited 2015 Nov 10]. Available from: https://www.hsph.harvard.edu/news/press-releases/diabetes-top-health-concern-for-latino-families/

10. Agardh E, Allebeck P, Hallqvist J, Moradi T, Sidorchuk A. Type 2 diabetes incidence and socio-economic position: a systematic review and meta-analysis. Int J Epidemiol. 2011 Jun 1;40(3):804–18.

11. Chaufan C, Davis M, Constantino S. The twin epidemics of poverty and diabetes: understanding diabetes disparities in a low-income Latino and immigrant neighborhood. J Community Health. 2011 Dec;36(6):1032–43.

12. Gaskin DJ, Thorpe Jr. RJ, McGinty EE, Bower K, Rohde C, Young JH, et al. Disparities in diabetes: the nexus of race, poverty, and place. Am J Public Health. 2014 Nov;104(11):2147–55.

13. Lysy Z, Booth GL, Shah BR, Austin PC, Luo J, Lipscombe LL. The impact of income on the incidence of diabetes: a population-based study. Diabetes Res Clin Pract. 2013 Mar 1;99(3):372–9.

14. Kaiser Family Foundation. Poverty Rate by Race/Ethnicity [Internet]. [cited 2022 Aug 29]. Available from: https://www.kff.org/other/state-indicator/poverty-rate-by-raceethnicity

15. US Census Bureau. 2009-2013 American Community Survey 5-Year Estimates [Internet]. [cited 2015 Jun 17]. Available from: https://www.census.gov/programs-surveys/acs/technical-documentation/table-and-geography-changes/2013/5-year.html

16. Community Preventive Services Task Force. Diabetes Prevention and Control: Combined Diet and Physical Activity Promotion Programs to Prevent Type 2 Diabetes Among People at Increased Risk [Internet]. Guide to Community Preventive Services. 2014 [cited 2015 Nov 10]. Available from: https://www.thecommunityguide.org/findings/diabetes-combined-diet-and-physical-activity-promotion-programs-prevent-type-2-diabetes.html

17. Guidelines for the Management of Diabetes Mellitus [Internet]. Michigan Quality Improvement Consortium; 2012 [cited 2015 Nov 10]. Available from: http://mqic.org/pdf/mqic_management_of_diabetes_mellitus_cpg.pdf

18. Haas L, Maryniuk M, Beck J, Cox CE, Duker P, Edwards L, et al. National standards for diabetes self-management education and support. Diabetes Care. 2014 Jan;36(Supplement 1):S100–108.

19. Knowler WC, Barrett-Connor E, Fowler SE, Hamman RF, Lachin JM, Walker EA, et al. Reduction in the incidence of type 2 diabetes with lifestyle intervention or metformin. The New England Journal of Medicine. 2002 Feb 7;346(6):393–403.

20. Barrera Jr M, Castro FG, Strycker LA, Toobert DJ. Cultural adaptations of behavioral health interventions: A progress report. Journal of Consulting and Clinical Psychology. 2013 Apr;81(2):196–205.

21. Hawthorne K, Robles Y, Cannings-John R, Edwards AGK. Culturally appropriate health education for Type 2 diabetes in ethnic minority groups: a systematic and narrative review of randomized controlled trials. Diabet Med. 2010 Jun;27(6):613–23.

22. Kong A, Tussing-Humphreys LM, Odoms-Young AM, Stolley MR, Fitzgibbon ML. Systematic review of behavioural interventions with culturally adapted strategies to improve diet and weight outcomes in African American women. Obes Rev. 2014 Oct;15(Supplement 4):62–92.

23. Lie DA, Lee-Rey E, Gomez A, Bereknyei S, Braddock CH. Does cultural competency training of health professionals improve patient outcomes? A systematic review and proposed algorithm for future research. J Gen Intern Med. 2011 Mar;26(3):317–25.

24. Nam S, Janson SL, Stotts NA, Chesla C, Kroon L. Effect of culturally tailored diabetes education in ethnic minorities with type 2 diabetes: a meta-analysis. J Cardiovasc Nurs. 2012 Nov 1;27(6):505–18.

25. Pottie K, Hadi A, Chen J, Welch V, Hawthorne K. Realist review to understand the efficacy of culturally appropriate diabetes education programmes. Diabetic Medicine. 2013 Sep;30(9):1017–25.

26. Ricci-Cabello I, Ruiz-Pérez I, Rojas-García A, Pastor G, Rodríguez-Barranco M, Gonçalves DC. Characteristics and effectiveness of diabetes self-management educational programs targeted to racial/ethnic minority groups: a systematic review, meta-analysis and meta-regression. BMC Endocr Disord. 2014 Dec;14(1):1–3.

27. Zeh P, Sandhu HK, Cannaby AM, Sturt JA. The impact of culturally competent diabetes care interventions for improving diabetes-related outcomes in ethnic minority groups: a systematic review. Diabet Med. 2012 Oct;29(10):1237–52.

28. Dauvrin M, Lorant V, d’Hoore W. Is the chronic care model integrated into research examining culturally competent interventions for ethnically diverse adults with Type 2 diabetes mellitus? A review. Evaluation & the Health Professions. 2015 Feb 4;38(4):435–63.

29. Page-Reeves J, Niforatos J, Mishra S, Regino L, Gingrich A, Bulten R. Health Disparity and Structural Violence: How Fear Undermines Health Among Immigrants at Risk for Diabetes. Journal of Health Disparities Research and Practice. 2013 Jan 1;6(2):30–47.

30. Page-Reeves J, Mishra SI, Niforatos J, Regino L, Bulten R. An Integrated Approach to Diabetes Prevention: Anthropology, Public Health, and Community Engagement. Qual Rep. 2013;18:1–22.

31. Page-Reeves J, Regino L, McGrew HC, Tellez M, Pedigo B, Overby A, et al. Collaboration and outside-the-box thinking to overcome training-related challenges for including patient Stakeholders as data collectors in a patient-engaged research project. Journal of Patient Experience. 2018 Jun;5(2):88–91.

32. Page-Reeves J, Regino L. Current Challenges of Community-University Health Research Partnerships and Concrete, Plain Language Strategies to Building Capacity. Anthropology in Action. 2018;In Press.

33. Page-Reeves J, Regino L, Tellez M, Pedigo B, Perez E. Engaging Latino Patients in Diabetes Research: What We Are Learning. Practicing Anthropology. 2018;40(3):35–9.

34. McGrew HC, Regino L, Bleecker M, Tellez M, Pedigo B, Guerrero D, et al. Training Patient Stakeholders Builds Community Capacity, Enhances Patient Engagement in Research. Journal of Community Engagement and Scholarship. 2020;13(1):99–106.

35. Funnell MM, Brown TL, Childs BP, Haas LB, Hosey GM, Jensen B, et al. National Standards for diabetes self-management education. Diabetes Care. 2011 Jan;34(Supplement 1):S89–96.

36. Stellefson M, Dipnarine K, Stopka C. The chronic care model and diabetes management in US primary care settings: a systematic review. Preventing Chronic Disease. 2013;10:E26.

37. American Diabetes Association. Recognized Education Programs– DiabetesPro [Internet]. [cited 2015 Nov 10]. Available from: https://professional.diabetes.org/diabetes-education

38. Parchman ML, Zeber JE, Palmer RF. Participatory decision making, patient activation, medication adherence, and intermediate clinical outcomes in type 2 diabetes: a STARNet study. Annals of Family Medicine. 2010 Sep 1;8(5):410–7.

39. Edelman D, Gierisch JM, McDuffie JR, Oddone E, Williams JW. Shared medical appointments for patients with diabetes mellitus: a systematic review. Journal of General Internal Medicine. 2015 Jan;30(1):99–106.

40. Clark L, Vincent D, Zimmer L, Sanchez J. Cultural values and political economic contexts of diabetes among low-income Mexican Americans. Journal of Transcultural Nursing. 2009 Oct;20(4):382–94.

41. Flores G. Culture and the patient-physician relationship: achieving cultural competency in health care. The Journal of Pediatrics. 2000 Jan 1;136(1):14–23.

42. Marin G, Marin BV. Research with Hispanic populations. Sage Publications, Inc; 1991.

43. Trickett EJ, Beehler S, Deutsch C, Green LW, Hawe P, McLeroy K, et al. Advancing the science of community-level interventions. American Journal of Public Health. 2011 Aug;101(8):1410–9.

44. Trickett EJ. Multilevel community-based culturally situated interventions and community impact: an ecological perspective. American Journal of Community Psychology. 2009 Jun;43:257–66.

45. Sixta CS, Ostwald S. Texas-Mexico border intervention by promotores for patients with type 2 diabetes. The Diabetes Educator. 2008 Mar;34(2):299–309.

46. Garcia AA, Villagomez ET, Brown SA, Kouzekanani K, Hanis CL. The Starr County Diabetes Education Study: development of the Spanish-language diabetes knowledge questionnaire. Diabetes Care. 2001 Jan 1;24(1):16–21.

47. Vincent D, Pasvogel A, Barrera L. A feasibility study of a culturally tailored diabetes intervention for Mexican Americans. Biological Research for Nursing. 2007 Oct;9(2):130–41.

48. Kim S, Love F, Quistberg DA, Shea JA. Association of health literacy with self-management behavior in patients with diabetes. Diabetes Care. 2004 Dec 1;27(12):2980–2.

49. Shah VO, Carroll C, Mals R, Ghahate D, Bobelu J, Sandy P, et al. A Home-Based Educational Intervention Improves Patient Activation Measures and Diabetes Health Indicators among Zuni Indians. PLoS ONE. 2015 May 8;10(5):e0125820.

50. Wolever RQ, Dreusicke M, Fikkan J, Hawkins TV, Yeung S, Wakefield J, et al. Integrative health coaching for patients with type 2 diabetes: a randomized clinical trial. The Diabetes Educator. 2010 Jul;36(4):629–39.

51. Druss BG, Zhao L, von Esenwein SA, Bona JR, Fricks L, Jenkins-Tucker S, et al. The Health and Recovery Peer (HARP) Program: A peer-led intervention to improve medical self-management for persons with serious mental illness. Schizophrenia Research. 2010 May 1;118:264–70.

52. Grønning K, Rannestad T, Skomsvoll JF, Rygg LØ, Steinsbekk A. Long-term effects of a nurse-led group and individual patient education programme for patients with chronic inflammatory polyarthritis - a randomised controlled trial. Journal of Clinical Nursing. 2014 Apr;23(7–8):1005–17.

53. Lorig K, Ritter PL, Villa FJ, Armas J. Community-based peer-led diabetes self-management: a randomized trial. The Diabetes Educator. 2009 Jul;35(4):641–51.

54. Huang FY, Chung H, Kroenke K, Delucchi KL, Spitzer RL. Using the Patient Health Questionnaire-9 to measure depression among racially and ethnically diverse primary care patients. Journal of General Internal Medicine. 2006 Jun;21(6):547–52.

55. Gilbody S, Richards D, Brealey S, Hewitt C. Screening for depression in medical settings with the Patient Health Questionnaire (PHQ): a diagnostic meta-analysis. Journal of General Internal Medicine. 2007 Nov;22(11):1596–602.

56. Reuland DS, Cherrington A, Watkins GS, Bradford DW, Blanco RA, Gaynes BN. Diagnostic accuracy of Spanish language depression-screening instruments. Annals of Family Medicine. 2009 Sep 1;7(5):455–62.

57. Looker HC, Knowler WC, Hanson RL. Changes in BMI and weight before and after the development of type 2 diabetes. Diabetes Care. 2001 Nov 1;24(11):1917–22.

58. Avery L, Flynn D, van Wersch A, Sniehotta FF, Trenell MI. Changing physical activity behavior in type 2 diabetes: a systematic review and meta-analysis of behavioral interventions. Diabetes Care. 2012 Dec 1;35(12):2681–9.

59. Bogner HR, Morales KH, de Vries HF, Cappola AR. Integrated management of type 2 diabetes mellitus and depression treatment to improve medication adherence: a randomized controlled trial. Annals of Family Medicine. 2012 Jan 1;10(1):15–22.

60. Lorig K, Ritter PL, Laurent DD, Plant K, Green M, Jernigan VBB, et al. Online diabetes self-management program: a randomized study. Diabetes Care. 2010 Jun 1;33(6):1275–81.

61. United States Census Bureau. American Community Survey Demographics and Housing Estimates 2012 [Internet]. [cited 2014 Sep 1]. Available from: https://www.census.gov/data/developers/data-sets/acs-5year/2012.html

62. Alderman SL, O’Donnell K, Monaco K. Bare Bones Budget: Measuring the Minimum Income Needed for the Bare Necessities of Families in New Mexico [Internet]. Albuquerque, NM: New Mexico Voices for Children; 2003. Available from: http://www.nmvoices.org/attachments/bbbfullreport.pdf

63. Page-Reeves J, Regino L, Erhardt EB, Murray-Krezan C, Pedigo B, Tellez M, et al. Patient-Centered Framework to Identify Culturally and Contextually Appropriate Options for Latinos with Diabetes from Low-Income Households. Patient-Centered Outcomes Research Institute; In Press.

64. Schiøtz ML, Bøgelund M, Almdal T, Jensen BB, Willaing I. Social support and self-management behaviour among patients with Type 2 diabetes. Diabetic Medicine: A Journal of the British Diabetic Association. 2012 May;29(5):654–61.

65. Rygg LØ, Rise MB, Grønning K, Steinsbekk A. Efficacy of ongoing group based diabetes self-management education for patients with type 2 diabetes mellitus. A randomised controlled trial. Patient Education and Counseling. 2012 Jan 1;86(1):98–105.

66. Hibbard JH, Mahoney ER, Stock R, Tusler M. Do increases in patient activation result in improved self-management behaviors? Health Services Research. 2007 Aug;42(4):1443–63.

67. Hibbard JH, Mahoney E. Toward a theory of patient and consumer activation. Patient Education and Counseling. 2010 Mar;78(3):377–81.

68. Hibbard JH. Community-based participation approaches and individual health activation. The Journal of Ambulatory Care Management. 2009 Oct 1;32(4):275–7.

69. Hendriks M, Rademakers J. Relationships between patient activation, disease-specific knowledge and health outcomes among people with diabetes; a survey study. Health Services Research. 2014 Dec;14:1–9.

70. Dixon A, Hibbard J, Tusler M. How do People with Different Levels of Activation Self-Manage their Chronic Conditions? The Patient: Patient-Centered Outcomes Research. 2009 Dec;2(4):257–68.

71. Hibbard JH, Mahoney ER, Stockard J, Tusler M. Development and testing of a short form of the patient activation measure. Health Services Research. 2005 Dec;40(6p1):1918–30.

72. Erhardt E, Murray-Krezan C, Regino L, Perez D, Bearer EL, Page-Reeves, Janet. Associations Between Depression and Diabetes Among Latinx Patients from Low-Income Households in New Mexico. Social Science & Medicine. 2023 Jan;320:115713.

73. Buuren S van, Groothuis-Oudshoorn K. mice: Multivariate Imputation by Chained Equations in R. Journal of Statistical Software. 2011 Dec 12;45(3):1–67.

74. Hammersley M. Questioning Qualitative Inquiry: Critical Essays. London: Sage; 2008.

75. Patient-Centered Outcomes Research Institute (PCORI). A Patient-Centered Framework to Test the Comparative Effectiveness of Culturally and Contextually Appropriate Program Options for Latinos with Diabetes from Low-Income Households [Internet]. 2016 [cited 2018 Sep 25]. Available from: https://www.pcori.org/research-results/2016/comparing-two-programs-help-latino-patients-low-income-households-manage-their-diabetes

76. Kroenke K, Spitzer RL, Williams JBW. The PHQ-9: Validity of a Brief Depression Severity Measure. Journal of General Internal Medicine. 2001 Sep 1;16(9):606–13.

77. Agency for Healthcare Research and Quality. About the CAHPS Cultural Competence Item Set [Internet]. 2012 [cited 2021 May 16]. Available from: https://archive.ahrq.gov/cahps/surveys-guidance/item-sets/cultural/2312_about_cultural_comp.pdf

78. Bates D, Mächler M, Bolker B, Walker S. Fitting Linear Mixed-Effects Models Using lme4. Journal of Statistical Software. 2015;67(1):1–48.

79. Kuznetsova A, Brockhoff PB, Christensen RHB. lmerTest Package: Tests in Linear Mixed Effects Models. Journal of Statistical Software. 2017 Dec 6;82(13):1–26.

80. Insignia Health. Patient Activation Measure (PAM)® [Internet]. Products. 2021 [cited 2016 Feb 2]. Available from: http://www.insigniahealth.com/products/pam-survey

81. Whittemore R. Culturally competent interventions for Hispanic adults with type 2 diabetes: a systematic review. Journal of Transcultural Nursing. 2007 Apr;18(2):157– 66.

82. Skyler JS, Bergenstal R, Bonow RO, Buse J, Deedwania P, Gale EA, et al. Intensive glycemic control and the prevention of cardiovascular events: implications of the ACCORD, ADVANCE, and VA diabetes trials: a position statement of the American Diabetes Association and a scientific statement of the American College of Cardiology Foundation and the American Heart Association. Journal of the American College of Cardiology. 2009 Jan 20;53(3):298–304.

83. Arnold LW, Wang Z. The HbA1c and all-cause mortality relationship in patients with type 2 diabetes is J-shaped: a meta-analysis of observational studies. The review of diabetic studies. 2014;11(2).

84. Diabetes Control and Complications Trial Research Group. The relationship of glycemic exposure (HbA1c) to the risk of development and progression of retinopathy in the diabetes control and complications trial. Diabetes. 1995 Aug 1;44(8):968–83.

85. Sherwani SI, Khan HA, Ekhzaimy A, Masood A, Sakharkar MK. Significance of HbA1c test in diagnosis and prognosis of diabetic patients. Biomarker insights. 2016 Jan;11:95–104.

86. Pankratz VS, Choi EE, Qeadan F, Ghahate D, Bobelu J, Nelson RG, et al. Diabetes status modifies the efficacy of home-based kidney care for Zuni Indians in a randomized controlled trial. Journal of Diabetes and its Complications. 2021 Feb 1;35(2):1–6.

87. Huang ES, Liu JY, Moffet HH, John PM, Karter AJ. Glycemic control, complications, and death in older diabetic patients: the diabetes and aging study. Diabetes care. 2011 Jun 1;34(6):1329–36.

88. Alameri M, Wafa W, AlTikriti A, Saadane I, Alkaf B, Lessan N. Incidence and Rate of Progression of Prediabetes to Type 2 Diabetes: A 10-Year Retrospective Study From UAE. Endocrine Practice. 2019 Apr 1;25:141–2.

89. Nelson RG, Pankratz VS, Ghahate DM, Bobelu J, Faber T, Shah VO. Home-based kidney care, patient activation, and risk factors for CKD progression in Zuni Indians: a randomized, controlled clinical trial. Clinical Journal of the American Society of Nephrology. 2018 Dec 7;13(12):1801–9.

90. Page-Reeves J, Murray-Krezan C, Regino L, Perez J, Bleecker M, Perez D, et al. A randomized control trial to test a peer support group approach for reducing social isolation and depression among female Mexican immigrants. BMC public health. 2021 Dec;21(1):1–18.

91. Page-Reeves J, Shrum S, Rohan-Minjares F, Thiedeman T, Perez J, Murrietta A, et al. Addressing Syndemic Health Disparities Among Latin Immigrants Using Peer Support. Journal of Racial and Ethnic Health Disparities. 2019 Apr;6(2):380–92.

92. Page-Reeves J, Moffett ML, Steimel L, Smith DT. The evolution of an innovative community-engaged health navigator program to address social determinants of health. Progress in Community Health Partnerships: Research, Education, and Action. 2016;10(4):603–10.

93. Page-Reeves J editor. Women Redefining the Experience of Food Insecurity: Life Off the Edge of the Table. Lexington Books; 2014.

94. Page-Reeves J. Conceptualizing Food Insecurity and Women’s Agency: A Synthetic Introduction. In: Page-Reeves J, editor. Women Redefining the Experience of Food Insecurity: Life Off the Edge of the Table. Lanham, Maryland: Lexington Books; 2014. p. 3–44.

95. Janet Page-Reeves, Amy Anixter Scott, Maurice Moffett, Veronica Apodaca, Vanessa Apodaca. “Is always that sense of wanting … never really being satisfied”: Women’s Quotidian Struggles With Food Insecurity in a Hispanic Community in New Mexico. Journal of Hunger & Environmental Nutrition. 2014 Apr 3;9(2):183–209.

